# Long-term perturbation of the peripheral immune system months after SARS-CoV-2 infection

**DOI:** 10.1101/2021.07.30.21261234

**Authors:** Feargal J. Ryan, Christopher M. Hope, Makutiro G. Masavuli, Miriam A. Lynn, Zelalem A. Mekonnen, Arthur Eng Lip Yeow, Pablo Garcia-Valtanen, Zahraa Al-Delfi, Jason Gummow, Catherine Ferguson, Stephanie O’Connor, Benjamin AJ Reddi, David Shaw, Chuan Kok-Lim, Jonathan M. Gleadle, Michael R. Beard, Simon C. Barry, Branka Grubor-Bauk, David J. Lynn

**Affiliations:** Precision Medicine Theme, South Australian Health and Medical Research Institute, Adelaide, SA 5001, Australia; Women’s and Children’s Health Network, North Adelaide, SA, Australia; Molecular Immunology, Robinson Research Institute, University of Adelaide, Adelaide, SA, Australia; Viral Immunology Group, Adelaide Medical School, University of Adelaide and Basil Hetzel Institute for Translational Health Research, Adelaide, SA, Australia; Gene Silencing and Expression Core Facility, Adelaide Health and Medical Sciences, Robinson Research Institute, University of Adelaide, Adelaide, SA, Australia; Infectious Diseases Department, Royal Adelaide Hospital, Central Adelaide Local Health Network, Adelaide, SA, Australia; Intensive Care Unit, Royal Adelaide Hospital, Central Adelaide Local Health Network and Adelaide Medical School, University of Adelaide, Adelaide, SA, Australia; Microbiology and Infectious Diseases Department, SA Pathology, Adelaide, SA, Australia; Department of Renal Medicine, Flinders Medical Centre, Flinders University, Bedford Park, SA, 5042, Australia; Flinders Health and Medical Research Institute, Flinders University, Bedford Park, SA 5042, Australia; Research Centre for Infectious Diseases, School of Biological Sciences, University of Adelaide, Adelaide, SA 5005, Australia

## Abstract

Increasing evidence suggests immune dysregulation in individuals recovering from SARS-CoV-2 infection. We have undertaken an integrated analysis of immune responses at a transcriptional, cellular, and serological level at 12, 16, and 24 weeks post-infection (wpi) in 69 individuals recovering from mild, moderate, severe, or critical COVID-19. Anti-Spike and anti-RBD IgG responses were largely stable up to 24wpi and correlated with disease severity. Deep immunophenotyping revealed significant differences in multiple innate (NK cells, LD neutrophils, CXCR3^+^ monocytes) and adaptive immune populations (T helper, T follicular helper and regulatory T cells) in COVID-19 convalescents compared to healthy controls, which were most strongly evident at 12 and 16wpi. RNA sequencing suggested ongoing immune and metabolic dysregulation in convalescents months after infection. Variation in the rate of recovery from infection at a cellular and transcriptional level may explain the persistence of symptoms associated with long COVID in some individuals.

## Introduction

Coronavirus Disease 2019 (COVID-19) is caused by the severe acute respiratory syndrome coronavirus 2 (SARS-CoV-2), a highly infectious respiratory virus, resulting in the ongoing global pandemic. COVID-19 usually presents as an asymptomatic or mild to moderate respiratory infection in previously healthy individuals with symptoms that include fever, cough, headache, fatigue, myalgia, diarrhea, and anosmia (1, 2). However, in older individuals or in those with prior co-morbidities such as obesity or cardiovascular disease, COVID-19 can quickly develop into a severe and life-threatening disease requiring urgent intensive care support. While the death toll from COVID-19 has been devasting (>4 million as of 9 July 2021 according to the Johns Hopkins University Coronavirus Resource Center (3)), the vast majority of those infected fortunately do recover. It is now increasingly clear however that recovered individuals, even those who had mild COVID-19, can suffer from persistent symptoms for many months after infection (4), which is popularly referred to as long COVID. For example, a cohort study of COVID-19 patients (median age 57) discharged from hospital in Wuhan, China, 6 months prior, reported that 63% of patients presented with fatigue or muscle weakness; 23% sleep difficulties; and 23% anxiety or depression (5). Individuals who were previously severely ill during their hospital stay have ongoing impaired pulmonary function and abnormal chest imaging. Similar reports continue to pour in from around the world (6–11). While the majority of these reports involve patients who were hospitalized with COVID-19, persistent, albeit milder and less frequent symptoms have also been reported in non-hospitalized individuals months after recovery (12). These reports resemble similar post-infectious syndromes after other infections, such as Ebola (13) and SARS-CoV-1 (14), and suggest that there may be a long-lasting dysregulation of the immune response in individuals recovering from COVID-19.

Flow cytometric analysis of peripheral blood samples collected from convalescents in the U.S. (median 29 days post-infection) has revealed altered frequencies of innate and adaptive immune cell population including CD4^+^ and CD8^+^ T cell activation and exhaustion marker expression in recovered individuals (15). A similar study in Singapore (median 34 days post-infection) found increased levels of circulating endothelial cells and effector T cells in those recovering from active disease (16). Single cell RNA sequencing (scRNA-Seq) of peripheral blood mononuclear cells (PMBC) from a small (n=10) cohort of patients that were 7-14 days post-recovery also found an increased ratio of classical CD14^+^ monocytes with high inflammatory gene expression, decreased CD4^+^ and CD8^+^ T cells and significantly increased plasma B cells (17). scRNA-Seq profiling of PBMC gene expression in a larger cohort of recovering individuals (n=95) found those with severe disease (n=36) had decreased plasmacytoid Dendritic Cells (pDCs) and increased levels of proliferative effector memory CD8^+^ T cells, relative to healthy controls (18). A potential limitation of this study, however, was that samples from recovered individuals were not collected at uniform timepoints during recovery, instead samples were collected between 9- and 126-days post-infection (on average

44.5 days). Longitudinal profiling of the transcriptome of PBMC collected from individuals (n=18) during treatment, convalescence, and recovery phases of infection (up to 10 weeks post-infection) revealed that recovery from COVID-19 was marked by decreased expression of genes involved in the interferon response, humoral immunity and increased signatures indicative of T-cell activation and differentiation (19). However, these responses were not compared with healthy controls. Another recent study longitudinally profiled immune cell populations and the blood transcriptome in >200 SARS-CoV-2 infected patients over 12 weeks from symptom onset to recovery (20). They compared the blood transcriptome in 2- time bins (0-24 and 25-48 days from symptom onset) and found substantial changes in immune cell populations and increased expression of genes involved in immunometabolism and inflammation, which persisted after infection.

Here, we have performed anti-receptor binding domain (RBD) and anti-Spike serology, comprehensive multi-parameter immunophenotyping, and transcriptome-wide RNA sequencing on blood collected from individuals recovering from mild/moderate or severe/critical COVID-19 at 12, 16, and 24 weeks after their first positive SARS-CoV-2 PCR test, as well as age-matched healthy controls (HCs). Our analyses reveal robust but heterogenous humoral immunity in convalescents until at least 6 months post-infection. Deep immunophenotyping highlighted profound changes in immune cell populations in COVID-19 convalescents compared with HCs, particularly at 12 and 16 weeks post-infection (wpi). Furthermore, RNA sequencing revealed significant changes in whole blood gene expression for up to 24 wpi, even in individuals that had mild disease without hospitalisation. These data suggest that SARS-CoV-2 infection leads to persistent changes to the peripheral immune system long after the infection is cleared which has important potential implications for understanding symptoms associated with long COVID. These changes to the peripheral immune system could have implications for how individuals recovering from infection respond to other challenges encountered in this period and persistent immune activation may also exacerbate other chronic conditions.

## Results

To assess the long-term effects of SARS-CoV-2 infection on the peripheral immune system, blood samples were collected from 69 recovering/convalescent COVID-19 indviduals at 12, 16 and 24 weeks (+/- 14 days) post-infection (wpi) **(Fig. 1A)**. Blood samples were also collected from n=14 seronegative HCs with no history of prior SARS-CoV-2 infection. COVID-19 convalescent individuals were classified according to the NIH classification of disease severity (21) as mild (n=50), moderate (n=6), severe (n=7) or critical (n=6) (**Table S1**). HCs were age- and sex-matched with mild/moderate convalescent individuals, however, as expected based on previous reports, severe/critical convalescent individuals were older and mostly male **(Fig. 1B-C)**. All samples in this study were collected in South Australia where early and strict international and interstate border control measures eliminated community transmission of the virus during the sample collection period (22). None of the participants had received a COVID-19 vaccine at the time of sample collection. This cohort was therefore uniquely placed for the assessment of immune responses in COVID-19 convalescents due a negligible risk of re-infection or changes induced by vaccination.

**Figure 1.**
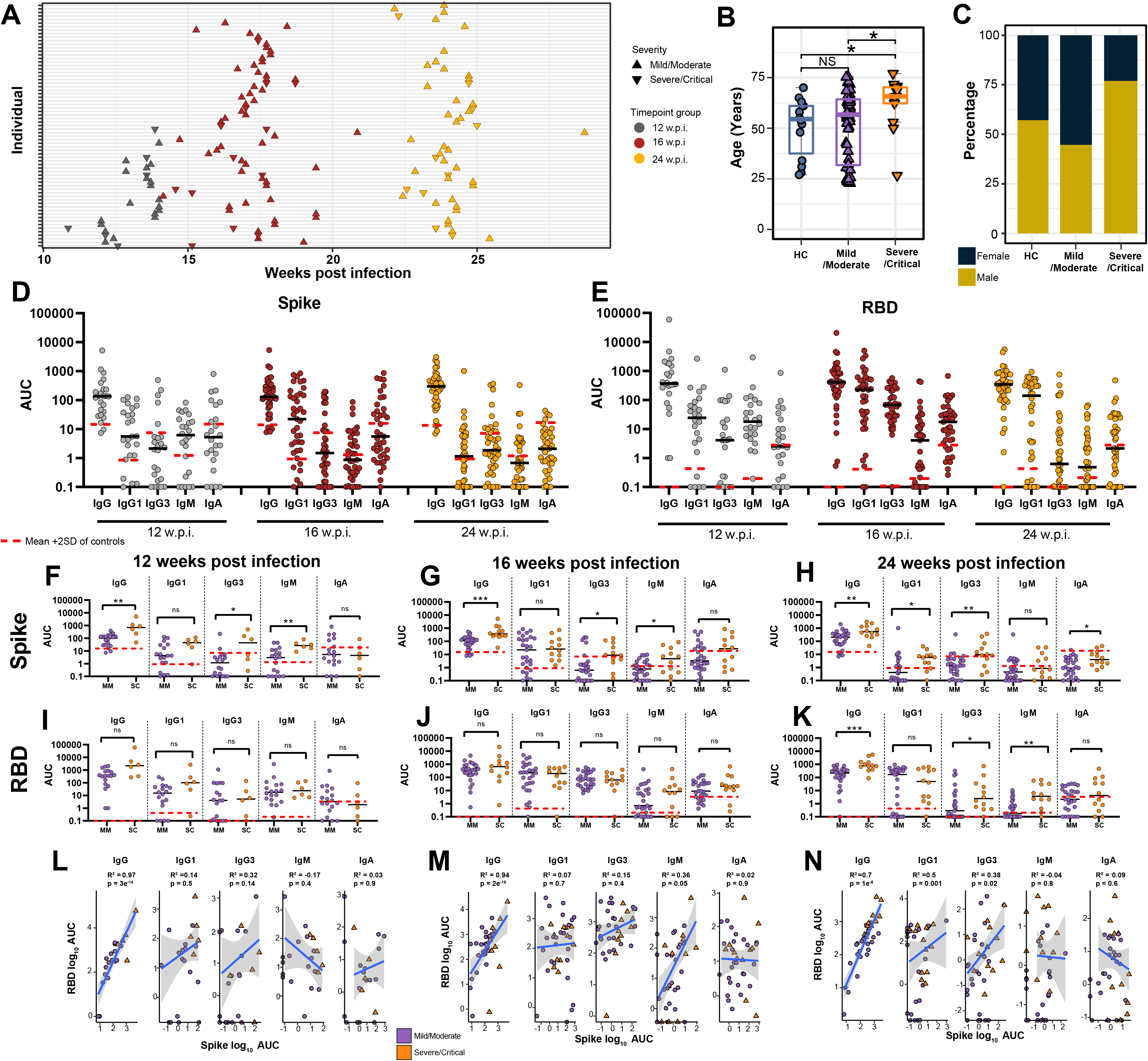
Anti-Spike and anti-RBD specific antibodies at 12, 16, and 24 weeks post-infection (w.p.i.). **(A)** Blood sample collection timepoints. **(B)** Age and **(C)** sex distribution of healthy controls (HC) in comparison to mild/moderate and severe/critical COVID-19 convalescents. **(D)** Anti-Spike and **(E)** anti-RBD specific IgG, IgG1, IgG3, IgM and IgA titres at 12, 16, and 24 w.p.i. End point titers are reported as area under the curve (AUC). The mean is denoted by the horizontal black lines. Seronegative samples were assigned a value of 0.1. Red dashed lines represent the mean AUC + 2 SD in HC for each isotype. **(F-K)** Antibody titres subdivided by disease severity. **(L-M)** Pearson correlations between anti-Spike and anti-RBD antibody subclass titres at each timepoint. Statistical significance was assessed in (B,F-K) using Wilcoxon Rank Sum Tests. ns= non-significant. * P < 0.05, ** P < 0.01, *** P < 0.001.

### COVID-19 convalescents display robust anti-Spike and anti-RBD antibody responses for at least 6 months post-infection

Anti-SARS-CoV-2 Spike and receptor binding domain (RBD) total IgG, IgG1, IgG3, IgM and IgA responses were evaluated in convalescent individuals at 12, 16, and 24 wpi **(Fig. 1D- E)**. The titres of Spike-specific IgG were diverse but largely stable over time **(Fig. S1A-C)**, although there was a trend for anti-Spike IgG1 titres to decline over time **(Fig. S1B)**. The seropositivity of Spike-specific serum IgM and IgA gradually diminished over time (**Fig. S1D-E**). Overall, the kinetics for anti-RBD antibodies were similar to those observed for anti-Spike antibodies **(Fig. S1F-J)**, though anti-RBD IgG3 and IgM appeared to decline more rapidly than anti-Spike antibodies. We also compared the levels of anti-Spike and anti-RBD circulating antibodies between individuals recovering from mild/moderate versus severe/critical COVID-19. Anti-Spike total IgG and IgG3 levels at 12, 16 and 24 wpi were significantly higher in severe/critical convalescents compared to those with previous mild/moderate disease **(Fig. 1F-H)**. Anti-Spike IgG1 and IgA levels were significantly higher in severe/critical convalescents at 24 wpi only (**Fig. 1H**). There was no detectable difference in RBD-specific antibody responses between individuals recovering from mild/moderate or severe/critical disease at 12 or 16 wpi (**Fig. 1I-J**). At 24 wpi severe/critical convalescent individuals maintained significantly higher anti-RBD IgG, IgG3 and IgM levels compared to individuals with previous mild/moderate COVID-19 disease (**Fig. 1K**). Anti-Spike and anti-RBD total IgG levels (but not other antibody subclasses) were significantly correlated at all timepoints (**Fig. 1L-N**). Anti-Spike and anti-RBD total IgG1 and IgG3 levels were significantly correlated at 24 wpi only. In summary, anti-Spike and anti-RBD antibody titers were generally positively correlated with COVID-19 disease severity, in accordance with previous observations (23–25).

### Deep immunophenotyping reveals persistent alterations in immune cell populations in COVID-19 convalescents up to 24 weeks post-infection

We used a multi-parameter flow cytometry approach to identify and enumerate ∼130 different immune cell sub-populations in samples collected from COVID-19 convalescent individuals at 12, 16 and 24 wpi and from HCs **(Table S2; Document S1)**. Our analysis included deep immunophenotyping of the CD4 and CD8 compartments, interrogating their maturation status, and in the CD4 compartment, interrogation of T helper (Th) lineage subsets, T regulatory (Treg) subsets, and T follicular helper (Tfh) subsets using a combination of chemokine receptor expression patterns to resolve Th lineages (Th1, 2, 17, 1/7, 9, 22, 2/22). Immune cell populations were first categorised into 10 major lineages (**Fig. 2A)**. Each cell type was further segregated based on functional marker characteristics including activation or maturation status. Differences in these major lineages, compared with HCs, were most strongly evident at 12 wpi but some populations were still significantly different at 24 wpi **(Fig. 2A-D, Table S2)**. While there was significant lymphopenia evident in convalescent individuals at 12 and 16 wpi **(Fig. 2E)**, CD3^+^ T cells were significantly increased at 12 wpi **(Fig. 2F)**. CD19^+^ B cells were also significantly increased at 12 and 16 wpi **(Fig. 2G)**. We also observed significantly increased CD38^+^CD27^+^ memory B cells at 16 wpi **(Fig. 2A)**. When interrogating CD4^+^ T cell maturation, we observed a significant reduction in both the CD4^+^ and CD8^+^ compartments at 12 and 16 wpi **(Fig. 2B-C)**. CD4^+^ effector memory (EM) pools were significantly reduced **(Fig. 2H)** and we also observed a significant reduction in migratory central memory (CM) CD4^+^ T cells, defined as CCR7^+^CD62L^-^, at all timepoints **(Fig. 2I)**.

**Figure 2.**
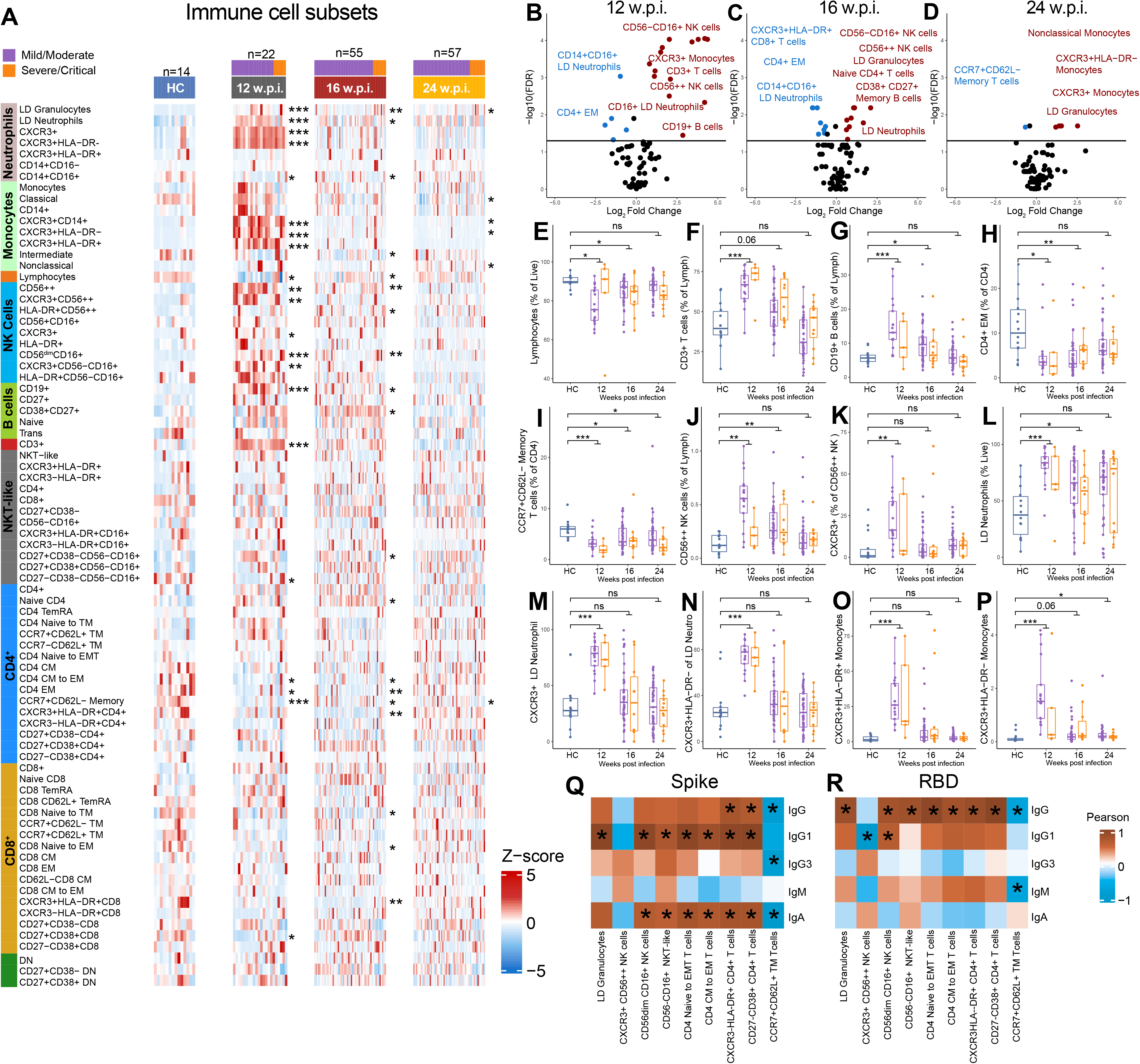
Flow cytometry analysis of major immune cell populations in peripheral blood mononuclear cells (PMBCs) collected from COVID-19 convalescents at 12, 16, and 24 weeks post-infection (w.p.i.) and from healthy controls (HC). **(A)** Heatmap representing the frequency of immune cell populations in HC and in convalescents. Brighter red color represents higher frequency. **(B-D)** Volcano plots of immune cell populations at each timepoint. Horizontal line represents FDR=0.05. Populations shown in red or blue were significantly (FDR < 0.05) increased or decreased (fold change > 1.5-fold), respectively, in convalescents. **(E-P)** The proportion of selected immune cell populations at 12, 16, and 24 w.p.i. compared to HC. Immune cell populations at 12 w.p.i. that were significantly correlated (FDR < 0.05 indicated by *) with **(Q)** anti-Spike or **(R)** anti-RBD antibody subclass titres at 24 w.p.i. The color intensity represents the strength of the correlation based on the Pearson correlation coefficient (red, positive correlation; blue, negative correlation). Only immune cell populations which were significantly correlated (with an absolute r^2^ > 0.7) with at least 1 anti-Spike or anti-RBD antibody subclass are shown. See Table S3 for all significant correlations. Statistical significance was assessed in (A-P) using Wilcoxon Rank Sum Tests. P values were adjusted for multiple testing using the Benjamini-Hochberg method. ns = non-significant. * FDR < 0.05, ** FDR < 0.01, *** FDR < 0.001.

The NK cell compartment was also altered in convalescents at 12 and 16 wpi **(Fig. 2A-C)** with CD56^++^ NK cells significantly elevated at 12 wpi whether enumerated as total **(Fig. 2J)** or tissue migratory (CXCR3^+^) **(Fig. 2K)**. We also observed a significant increase in total granulocytes at all 3 timepoints post-infection **(Fig. 2A-D)**, and this was also observed for low density (LD) neutrophils at 12 and 16 wpi **(Fig. 2L)**. CXCR3^+^ LD neutrophils, which are actively recruited to sites of tissue damage (26), were elevated in convalescents at 12 wpi but returned to baseline by 16 wpi **(Fig. 2M-N)**. Interestingly, CD14^+^CD16^+^ neutrophils were significantly decreased at 12 and 16 wpi (**Fig. 2A)**. While total monocyte proportions were not significantly altered, two subsets of tissue-homing CXCR3^+^ monocytes (HLA-DR^+^, activated antigen-presenting proinflammatory monocytes and HLA-DR^-^, regulatory monocytes) were significantly increased in convalescent individuals at 12 wpi **(Fig. 2O-P)**.

We also investigated differences in immune cell populations between mild/moderate and severe/critical convalescents, however, after correction for multiple testing there were no statistically significant differences **(Table S2)**, most likely due to the small sample size of severe/critical convalescent samples, particularly at 12 wpi.

Next, we assessed correlations between immune cell populations (at 12, 16 or 24 wpi) and both anti-Spike and anti-RBD IgG, IgM, and IgA responses at 24 wpi **(Fig. 2Q-R, Table S3)**. Significant positive correlations were observed between the frequency of granulocytes, CD16^+^ NK and NKT-like cells at 12 wpi and anti-Spike IgG1 and anti-RBD IgG titres at 24wpi. These data may be reflective of the correlation between disease severity and antibody responses. Previous work has suggested an association between increased percentage of neutrophils and lower anti-RBD IgG responses (27), which we did not detect in our analysis (**Table S3**). Components of the CD4 compartment were also significantly associated with anti-Spike IgG1 and anti-RBD IgG titres at 24 wpi. For example, there was a positive correlation between the proportion of CD4^+^ cells in transition from naïve to CM, CM to EM CD4^+^ T cells, and activated (HLA-DR^+^ or CD38^+^) CD4^+^ T cells and anti-Spike and anti-RBD IgG/G1 titres at 24 wpi, suggesting each of these CD4 populations might contribute to robust T cell help. Significant correlations between immune cell populations at 16 and 24 wpi and anti-Spike or anti-RBD antibody responses were also observed **(Table S3)**.

To interrogate CD4 Th responses in more depth, we applied a chemokine receptor-based gating strategy to characterise the Th effector phenotypes in both Th and Tfh subsets (28, 29). We also used CD45RO^+^ and CD62L^+^ staining as a marker of T cell memory formation in the Th subsets. In addition, we applied the same strategy to T regulatory (Treg) subsets, which are functionally paired with their Th and Tfh counterparts *in vivo* (28). Th and Tfh lineages were categorised into 8 functional subsets **(Fig. 3A)** and significant differences were observed for multiple subsets in COVID-19 convalescents **(Fig. 3B-D, Table S2)**. We observed a significant decrease in Th9 cells at all timepoints **(Fig. 3E)**. There was also a significant increase in Th2/22 cells at 16 wpi **(Fig. 3A)**. This may reflect a failure to mount a normal tissue repair response in the mucosa in the lung, as the Th9 and Th22 family are predicted to home to epithelial mucosa (30, 31). Alternatively, these subsets may be underrepresented in the circulation as they have transmigrated to sites of damage. In examining the formation of Th cell memory, we observed that while the proportion of Th17 and Th22 cells was not significantly different between groups, there was an increased proportion of Th17 and Th22 CM cells at all timepoints **(Fig. 3F-G)**. This may indicate a role for these mature subsets in antiviral responses. In addition, there was evidence of increased formation of Th2/22 memory at 12 wpi **(Fig. 3H)**, suggesting establishment of memory focused on tissue repair (32). In the Tfh compartment, we observed significant differences in Tfh1, 9, 22 and 2/22 cells at different timepoints post-infection **(Fig. 3A)**, with Tfh1 cells significantly elevated in convalescents at 12 and 16 wpi **(Fig. 3A)**.

**Figure 3.**
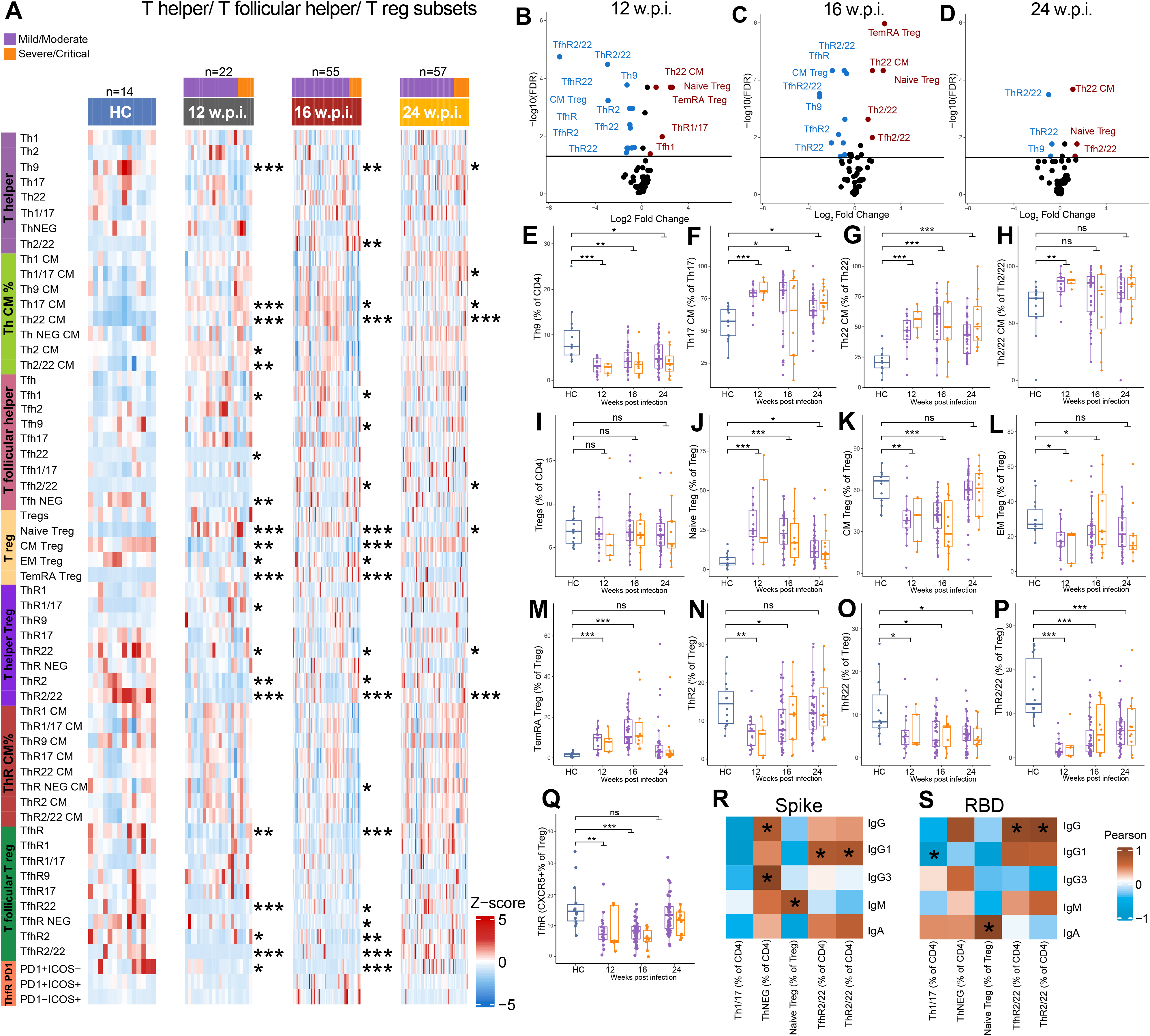
Flow cytometry analysis of T helper (Th), T follicular helper (Tfh), and T regulatory cell (Treg) populations in peripheral blood mononuclear cells (PMBCs) collected from COVID-19 convalescents at 12, 16, and 24 weeks post-infection (w.p.i.) and from healthy controls (HC). **(A)** Heatmap representing the frequency of immune cell populations in HC and in convalescents. Brighter red color represents higher frequency. **(B-D)** Volcano plots of immune cell populations at each timepoint. Populations shown in red or blue were significantly (FDR < 0.05) increased or decreased (fold change > 1.5-fold), respectively, in convalescents. **(E-Q)** The proportion of selected immune cell populations at 12, 16, and 24 w.p.i. compared to HCs. Th, Tfh, and Treg populations at 12 w.p.i. that were significantly correlated (FDR < 0.05; indicated by *) with **(Q)** anti-Spike or **(R)** anti-RBD antibody subclass titres at 24 w.p.i. The color intensity represents the strength of the correlation based on the Pearson correlation coefficient (red, positive correlation; blue, negative correlation). Only immune cell populations which were significantly correlated (with an absolute r^2^ > 0.6) with at least 1 anti-Spike or anti-RBD antibody subclass are shown. See Table S3 for all significant correlations. Statistical significance was assessed in (A-P) using Wilcoxon Rank Sum Tests. P values were adjusted for multiple testing using the Benjamini-Hochberg method. ns = non-significant. * FDR < 0.05, ** FDR < 0.01, *** FDR < 0.001.

As with CD8^+^ and CD4^+^ effector T cells, Tregs segregate into naïve and mature populations depending on antigen exposure. While we found no difference in total Tregs (**Fig. 3I**), we observed a significant increase in naïve Tregs at all timepoints post-infection **(Fig. 3J)**, accompanied by a significant decrease in CM and EM Tregs at 12 and 16 wpi, and a significant increase in TEMRA Tregs (effector memory with acquired CD45RA) at 12 and 16 wpi **(Fig. 3K-M)**. These data suggest either a block in maturation, or an increase in formation of naïve Treg cells in convalescents. The dual role of Treg cells in immune suppression and tissue repair suggest the potential for more than one mechanism of action in recovering individuals, so we examined functionally paired helper lineages in the Treg compartment, as they are likely to respond to the same pathogen-triggered homing cues as their Th effector counterparts. We observed a significant decrease in the proportion of ThR2 Tregs at 12 and 16 wpi, and a significant decrease in ThR22 and ThR2/22 Tregs at all timepoints **(Fig. 3N-P)**, suggesting a block in commitment of theses lineages. Finally, we also examined the follicular regulatory T cell lineages (TfhR), as they serve a similar regulatory role in germinal centres, controlling Tfh function and B cell help. We observed a significant decrease in total TfhR at 12 and 16 wpi **(Fig. 3Q)** suggesting that follicular help is less restrained by TfhR in individuals recovering from COVID-19. Specifically, TfhR2, 22 and 2/22 subsets were all significantly reduced at 12 and 16 wpi, but returned to baseline by 24 wpi. This is consistent with the regulatory follicular arm licencing a Tfhl response early in infection, but later, shaped by the tissue location of the virus, skewing to Tfh2/22 driven B cell help in germinal centres, both of which are required to drive an effective anti-viral B cell response.

We also sought to determine links between T cell help and antibody responses to COVID-19, given that priming and durable immunity are underpinned by the interaction of T and B cells. To do this, we performed a correlation analysis between CD4^+^ T cell subsets at 12, 16 and 24 wpi and antibody responses at 24 wpi **(Fig. 3R-S, Table S3)**. We observed a number of interesting statistically significant correlations. For example, we observed a significant positive correlation between anti-Spike IgG1 levels and both ThR2/22 and TfhR2/22 subsets, suggesting that the effector function of this epithelial tissue homing lineage may regulate antibody responses. Similar correlations between these subsets and anti-RBD IgG responses were also evident.

### Whole blood RNA sequencing reveals significant perturbations to gene expression in COVID-19 convalescents until at least 6 months post-infection

To assess the potential long-term effects of SARS-CoV-2 infection on the peripheral blood transcriptome, total RNA sequencing was performed on 138 blood samples collected from individuals recovering from mild (n=47), moderate (n=6), severe (n=7) or critical (n=6) COVID-19 at 12, 16 and 24 wpi **(Fig. 1A)**. RNA sequencing was also performed on blood collected from age-matched HCs (n=14) with negative serology for the SARS-CoV-2 Spike and RBD proteins. Approximately 9 billion 2×150bp read pairs (mean 68.2 million per sample) were sequenced (**Table S4**).

After adjusting for sex and batch effects, MDS analysis of the gene expression data revealed a clear separation between HCs and convalescent individuals at each timepoint (**Fig. 4A-C**). Consistent with these data, differential gene expression analysis identified >950 genes that were significantly (FDR < 0.05, fold change >1.25) differentially expressed (738 up- regulated genes; 230 down-regulated) in convalescent individuals at 12 wpi compared to HCs **(Fig. 4D, Table S4)**. Fewer differentially expressed genes (DEGs) were identified at 16 and 24 wpi, but there were still >250 DEGs identified at 24 wpi (**Fig. 4D, Table S4**).

**Figure 4:**
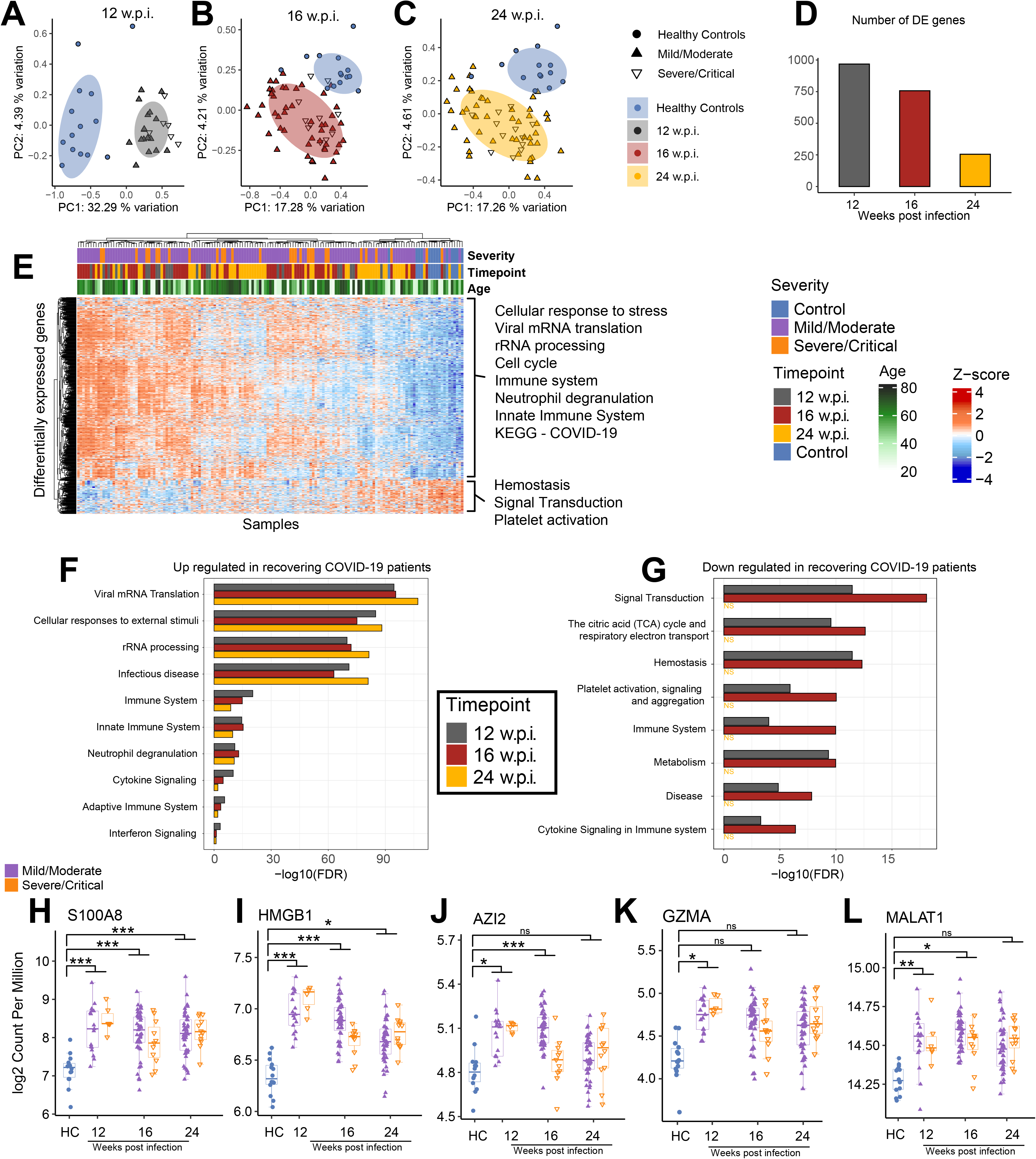
RNA-Seq was used to profile gene expression in peripheral whole blood samples collected from COVID-19 convalescents at 12, 16, and 24 weeks post infection (w.p.i.) and from healthy controls (HC). (**A-C**) Multidimensional scaling (MDS) analysis of RNA-seq gene expression data at 12, 16, and 24 w.p.i. compared to HC. (**D**) The number of differentially expressed (DE) genes (FDR < 0.05 and fold change >1.25-fold) identified at each timepoint. (**E**) Heatmap showing the expression of DE genes in each sample. Data were adjusted for sex and batch effects prior to MDS analysis and visualisation of the heatmap. (**F-G**) Selected REACTOME pathways enriched among **(F)** up-regulated and **(G)** down- regulated genes at each timepoint. See Table S4 for all enriched pathways. (**H-L**) The expression of selected genes in convalescents at 12, 16, and 24 w.p.i. compared to HC. Statistical significance comparing all convalescents to HC was assessed in (H-L) using EdgeR. P values were adjusted for multiple testing using the Benjamini-Hochberg method. ns = non-significant. * FDR < 0.05, ** FDR < 0.01, *** FDR < 0.001.

Unsupervised hierarchical clustering analysis of DEGs did not reveal an obvious clustering by disease severity, suggesting that even individuals with mild COVID-19 have long-lasting changes to their blood transcriptome (**Fig. 4E**). There was a tendency for samples from the earlier timepoints to cluster together, consistent with a decrease in the number of DEGs over time, but clearly there was a spectrum in the recovery in gene expression among convalescent individuals, with some recovering more quickly (clustering with HCs).

Pathway and Gene Ontology (GO) analysis revealed a very strong enrichment for pathways related to transcription, translation, and ribosome biosynthesis among genes up-regulated in convalescents, at all 3 timepoints (**Fig. 4E-F, Table S4**). In many cases these signatures were predominantly driven by the up-regulation of ribosomal RNA (rRNA) genes. Viral polypeptide synthesis is reliant upon host ribosomes and many viruses have been reported to stimulate rRNA synthesis upon infection (33, 34), although the SARS-CoV-2 Nsp1 protein has been shown to act a strong inhibitor of translation (35). Interestingly, a recent study has surprisingly shown that rRNA accumulation positively regulates antiviral innate immune responses against human cytomegalovirus infection (36), raising the possibility that the continued up-regulation of rRNAs in individuals recovering from COVID-19 is a cellular defence mechanism. Consistent with this, the Reactome pathway “innate immune system” was significantly enriched among genes up-regulated in convalescents (**Fig. 4E-F, Table S4)**. Other statistically enriched pathways among up-regulated genes included neutrophil degranulation, antimicrobial peptides, immune system, pathways related to other viral infections, cell cycle related pathways, and pathways related to the citric acid (TCA) cycle and respiratory electron transport/oxidative phosphorylation (**Table S4)**.

Among down-regulated genes at 12 and 16 wpi there was a strong enrichment for metabolic related pathways such as oxidative phosphorylation as well as pathways related to platelet activation, signaling and aggregation (**Fig. 4G**, **Table S4**). Platelet aggregation has previously been identified as a marker of severe SARS-CoV-2 infection (37), so it is interesting that genes involved in this process appear to be down-regulated in recovering individuals **(Fig. S2A, Table S4**). Interestingly, we identified oxidative phosphorylation to be enriched among up-regulated genes as well as down-regulated genes. Increased expression of genes involved in oxidative phosphorylation has recently been reported in another study assessing COVID-19 convalescents (20). Further examination of our data revealed that down- regulated oxidative phosphorylation genes were encoded by the mitochondria, whereas up- regulated ones were nuclear encoded (**Fig. S2B**). Differential expression of nuclear versus mitochondrially encoded oxidative phosphorylation genes has been reported in a number of other contexts (38). As mentioned, at 24 wpi there were considerably fewer DEGs (∼250) in convalescents compared to HCs, consistent with this, only one pathway, “complement activation”, was identified as being enriched among genes down-regulated at 24 wpi **(Table S4)**.

Many of the most strongly up-regulated genes in COVID-19 convalescents encoded known biomarkers of inflammation and innate immunity including S100 calcium-binding protein A8 (S100A8), and high-mobility group protein 1 (HMGB1), 5-azacytidine induced 2 (AZI2), and granzyme A (GZMA) (**Fig. 4H-K**). As we performed total RNA sequencing we were also able to identify many differentially expressed long-non-coding RNAs (**Table S4**) including metastasis-associated lung adenocarcinoma transcript 1 (MALAT1) (**Fig. 4L**), which has been found up-regulated in response to flavivirus and SARS-CoV-2 infection (39, 40) and is an important regulator of immunity and the cell cycle (41, 42).

As detailed above, flow cytometry analysis revealed significant changes in the proportion of multiple immune cell populations in convalescent individuals compared with HCs **(Fig. 2-3)**. As we performed RNAseq on whole blood samples, it was therefore possible that the differences we observed in the transcriptome of recovering individuals simply reflected changes in immune cell populations, rather than differences in gene expression. To assess this, we repeated the differential expression analysis multiple times, each time adjusting for changes in a major immune cell population. We found that accounting for changes in specific immune cell populations in our differential gene expression analysis models resulted in a decrease in the number of genes identified as differentially expressed (mean reduction in the number of genes with FDR < 0.05 was 50.47%), however, the statistical enrichment of immune, rRNA processing, cell cycle and transcription/translation signatures identified among DEGs was robust to correction for differences in the proportion of any specific immune cell populations (**Fig. S3, Table S4**). These data indicate that the long-term perturbation of the blood transcriptome that we observe in convalescents compared to HCs is not solely explained by changes in the frequency of any single immune cell population.

### Blood transcriptional module analysis highlights variable rates of recovery in the transcriptome of COVID-19 convalescents and correlations with antibody responses

We next sought to investigate individual-specific transcriptional changes in COVID-19 convalescents using pre-defined blood transcriptional modules (BTMs) (43). To do this, we used Gene Set Variation Analysis (GSVA) (44) to reduce variation captured across >20,000 genes in our gene expression data to an “activity score” for 256 BTMs in each individual (**Fig. 5A and Table S5**). Using *limma* we identified 80 of these BTMs that were differentially active in convalescents **(Table S5)**. The annotation of these BTMs was broadly consistent with our pathway analysis identifying multiple modules related to transcription/translation, the cell cycle and specific immune cell populations and pathways as being significantly enriched in convalescents (**Fig. 5A, Table S5**). Interestingly, this analysis highlighted that while the proportion of recovering COVID-19 convalescents with ‘healthy-like’ BTM activity increased over time (consistent with a recovery to baseline over time), there were still a subset of convalescents with persistent transcriptional dysregulation at 24 wpi (red and blue modules in **Fig. 5A**).

**Figure 5:**
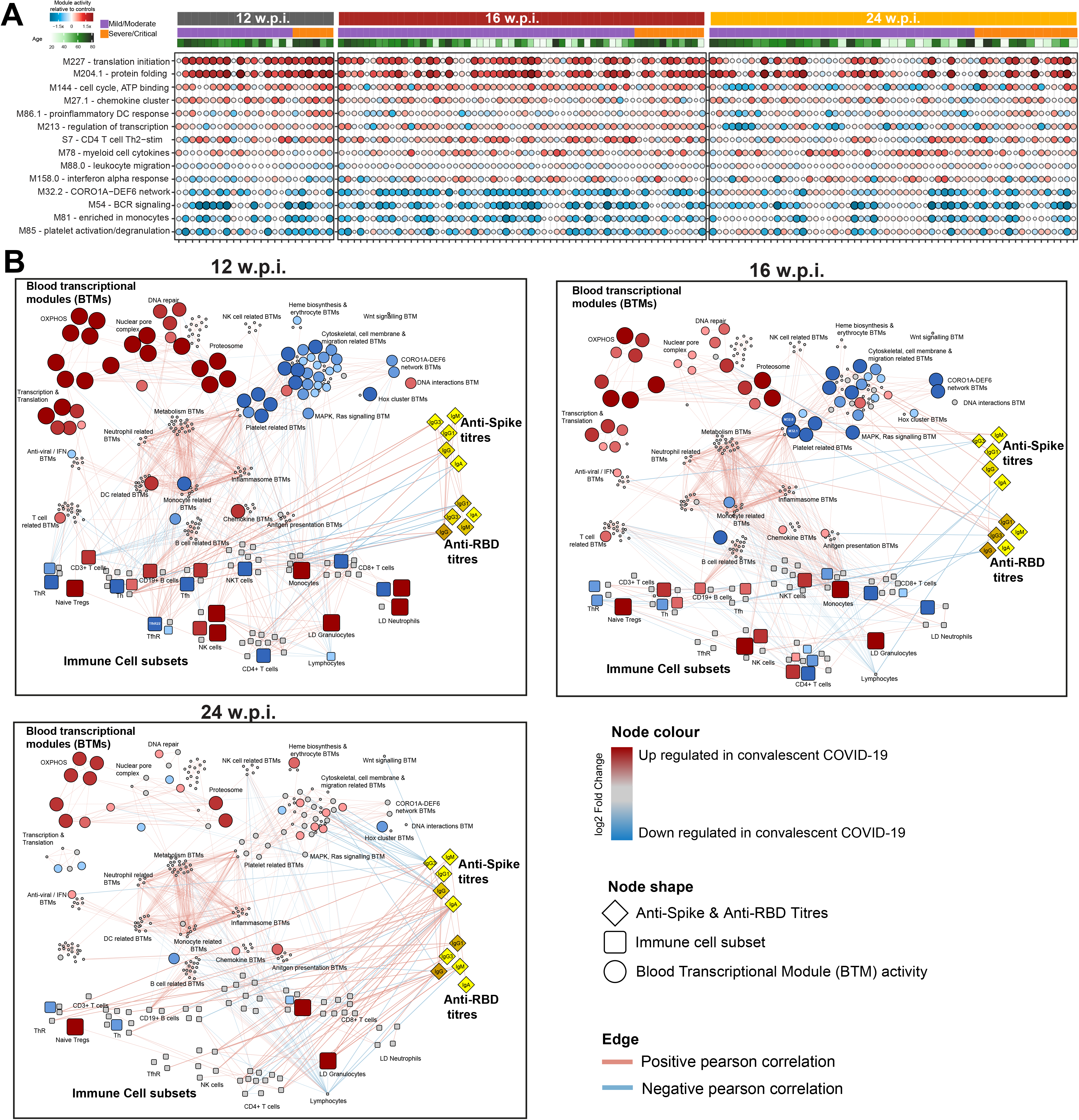
Integrated network analysis of correlations between blood transcriptional modules (BTMs), the frequency of immune cell populations assessed by flow cytometry and anti-Spike and anti-RBD antibody titres. **(A)** Selected BTMs identified to be differentially active in COVID-19 convalescents. Each circle represents the activity of that BTM in a specific convalescent individual. Darker red indicates increased BTM activity relative to healthy control (HC); Darker blue decreased. The size of the circle is proportionate to BTM activity relative to HC. **(B)** Network showing Pearson correlations (as edges) between BTMs, immune cell populations and serology data. Red and blue edges indicate positive and negative correlations, respectively. BTM-BTM correlations were determined across all timepoints. Only those with r^2^ >0.7 and FDR < 0.05 are shown. Correlations between BTMs, immune cell populations and antibody titres were determined at each timepoint. Only those with FDR < 0.05 at a specific timepoint are shown. Node sizes and colours are scaled relative to HC. Red and blue nodes indicate increased and decreased values, respectively, relative to HC. Grey nodes were not significantly altered in convalescents. The network was visualised using Cytoscape v3.8.1.

Finally, we undertook a systems-level integration of BTM activity scores, anti-Spike and anti-RBD antibody data, and flow cytometry data at 12, 16 and 24 wpi (**Fig. 5B, Table S5**). To do this, we constructed a network of significant correlations between BTMs, antibody titres and the frequency of immune cell populations in each individual. Many BTMs, including those differentially active in convalescents, were strongly correlated with each other. For example, monocyte, DC, neutrophil, and inflammasome related BTMs were strongly correlated with each other and, interestingly, with metabolism related BTMs **(Fig. 5B)**. DC related BTMs also correlated with antiviral and interferon related BTMs. At 12 wpi there were also significant correlations identified between BTMs and >30 different immune cell populations. For example, the proportion of CD19^+^ B cells, CD3^+^ T cells and NK cells were strongly correlated with the activity scores of BTMs independently annotated to be related to these cell types (**Fig. 5B, Table S5**). Multiple different immune-related BTMs were also strongly positively correlated with the proportion of Tfh22-like Tregs at 12wpi. At 16 wpi, BTMs correlated with fewer immune cell populations, however, multiple strong correlations were identified between BTMs and monocytes, B cells, and lymphocytes at this timepoint. Similar correlations were also identified at 24 wpi. Despite the strong correlations between many different BTMs and immune cell populations, interestingly, there were relatively few significant correlations between immune cell populations and differentially active BTMs, particularly at 16 and 24 wpi. These data suggest that the majority of differentially active BTMs we observed in convalescent individuals are not explained by differences in the frequency of immune cell populations in these individuals. For example, oxidative phosphorylation related BTMs were differentially active at all timepoints, however, these BTMs were not significantly correlated with any immune cell population. Anti-Spike and/or anti-RBD antibody titres were also significantly correlated with BTMs at 16 and 24 wpi, but not 12 wpi (**Fig. 5B, Table S5**). Many of the BTMs that were correlated with antibody titres were down-regulated in convalescents. For example, two platelet activation BTMs (M32.0 and M32.1) were significantly correlated with anti-Spike IgM responses at 16 wpi, while multiple cell adhesion related BTMs were significantly negatively correlated with anti-Spike IgG responses at 24 wpi. We also identified that multiple different immune cell populations that correlated with antibody titres at each timepoint. These relationships were particularly evident at 24 wpi. For example, the proportion of LD granulocytes, CD16^+^ NK cells and CCR7^-^CD62L^+^ transitional memory T cells were significantly positively correlated with anti-Spike and anti-RBD IgG titres at 24 wpi. In summary, our integrated network analysis reveals a complex interplay of relationships between circulating immune cell populations, transcriptional dysregulation, and humoral immune responses in COVID-19 convalescent patients and provides a resource for further exploration and investigation of these relationships.

## Discussion

Recovery from SARS-CoV-2 infection is frequently associated with persistent symptoms months after infection including fatigue, muscle weakness, sleep impairment and anxiety or depression (4, 5, 45). These data suggest ongoing immune dysregulation in COVID-19 convalescents which has been supported by several recent studies profiling the immune system in individuals recovering from COVID-19 using multi-parameter flow cytometry, bulk and single-cell transcriptomics, and other approaches (15, 16, 46–49). Our study extends on these recently published studies, which have mostly assessed immune responses at 2-12 weeks post-infection. Here, we report an integrated analysis of immune responses at a transcriptional, cellular and serological level, in individuals recovering from mild/moderate or severe/critical COVID-19 at 12, 16, and 24 weeks post-infection, in comparison to age-matched HCs.

Anti-Spike and anti-RBD serology data demonstrated heterogeneity of antibody responses to SARS-CoV-2 consistent with previously published reports showing long-lasting IgG and IgG1 antibody responses to at least 6 months post-infection which were correlated with disease severity (23, 50, 51). Our cohort is particularly well-suited to the assessment of the durability of antibody responses due to the negligible risks of re-infection in South Australia where, due to strict border restrictions and public health measures, community transmission was eliminated during the sample collection period. Despite the anticipated decay in IgA and IgM (52–54) a large percentage of convalescents remained seropositive for both RBD- and Spike-specific Ig (all isotypes) for the duration of the study. This decay was less pronounced at 24 wpi in the severe COVID-19 convalescents compared to the mild cohort, with significant differences in RBD-specific IgM and IgG3 isotypes between the two groups. Recently, declining levels of SARS-CoV-2 Spike-specific IgM in mild COVID-19 convalescents were found to strongly correlate with serum virus neutralisation activity (55), findings that were further confirmed in experiments with purified IgM fractions and IgM-depleted sera from similar patients (27, 56). In COVID-19 convalescents, IgM, similarly to IgG1, preferentially targets the S1 domain of the Spike protein (57), the region that contains the RBD and N-terminus domains and the target of most neutralising antibodies and regions of high interest for developing passive immunotherapies to deal with new SARS-CoV-2 variants of concern (58). Conversely, less abundant SARS-CoV-2-specific IgG3, targets the S2 domain more efficiently (57), which suggests that its ability to neutralise the virus is, by comparison, reduced. Yet, S2 contains the sequences that allow SARS-CoV-2 membrane fusion with the cell host membrane, a key step in virus entry (2). In fact, the ability of antibodies targeting S2 regions involved in membrane fusion to block Spike protein-mediated cell-cell fusion has been confirmed experimentally (59). In the future it will be necessary to elucidate the particular roles of IgM and IgG3 in neutralising SARS-CoV-2 but, perhaps too, blocking virus infection by other mechanisms such as blockade of membrane fusogenic regions of the Spike protein. This will provide further insights into the overall importance of specific Ig isotypes in determining disease severity and outcomes.

In addition to our serological analysis of COVID-19 convalescents, we extensively and longitudinally profiled immune cell populations in the same individuals using a multi-panel approach that enabled the identification and enumeration of ∼130 different sub-populations including deep phenotyping of the CD4 and CD8 compartments. Differences in immune cell populations compared with HCs were most strongly evident at 12 wpi, but some populations were still significantly different at 24 wpi. CD56^++^ NK cells, granulocytes, LD neutrophils and tissue-homing CXCR3^+^ monocytes were significantly increased in convalescents at 12 wpi. Many of these changes persisted until at least 16 or 24 weeks. Consistent with our data, increased NK cells (46) and granulocytes (49) have been reported in other cohorts of convalescents and scRNAseq has revealed that increased non-classical monocytes are associated with more severe disease during active infection (60). In contrast to our study, a study of 109 Austrian convalescents at 10 weeks post-infection, did not find neutrophils, monocytes, CD3^+^ T cells, CD56^+^ NK cells or CD19^+^ B cells to be significantly different in convalescents (49). Other studies have also reported significant decreases in the frequencies of invariant NKT and NKT-like cells (47), which we and others (20) did not observe.

Several previous studies have reported that T and B cell activation/exhaustion markers remain elevated following SARS-CoV-2 infection (15). Furthermore, CD4^+^ and CD8^+^ EM T cells have been reported to be significantly higher in convalescents at 10 wpi (49). Consistent with reports in active infection and convalescence (15), convalescent individuals in our study had lymphopenia until at least 16 wpi, however, CD3^+^ T cells were significantly increased at 12 wpi. We also observed significantly increased CD19^+^ B cells at 12 and 16 wpi and CD38^+^CD27^+^ memory B cells at 16 wpi in convalescents. Recent studies have shown that increased activation and exhaustion of memory B cells observed during COVID-19 correlates with CD4^+^ T cell functions (61), and consistent with this we observed reduced CD4^+^ EM cell proportions in COVID-19 convalescents at 12 wpi. We were particularly interested in the role of regulatory T cells (Tregs) in COVID-19, as there have been conflicting reports of Tregs being either increased or decreased in convalescents. Significantly increased Foxp3^+^ Tregs were observed in 49 convalescents from Wuhan at ∼112 days post-recovery (47), however, another study observed that CD25^+^Foxp3^+^ Tregs were significantly reduced 10 weeks after COVID-19 (49). We observed no significant difference in the total (CD4^+^CD25^+^CD127^low^) Treg pool at any timepoint, but when we interrogated Tregs for their memory/maturation status, we observed that the naïve and TEMRA Treg compartment was significantly expanded at 12 and 16 wpi, while EM and CM Tregs were significantly reduced, mirroring a similar reduction in the proportion of CD4^+^ EM and CM pools at 12 and 16 wpi.

Interestingly, a number of the Th lineage subsets including Th2, Th22, Th2/22, and Th17 had an increased proportions of CM vs EM, revealing subtle skewing of the Th memory formation. The expansion of naïve Tregs could be an attempt to restore the balance in the Treg pool in the face of both inflammation and tissue damage, which is supported by emerging evidence of a dual role for Tregs in supressing immune responses and promoting tissue repair (62). Increased TEMRA Tregs, which are often associated with exhaustion, but are in fact a poly-functional effector Treg population with characteristics of cytotoxic cells, migratory T cells and tissue repair cells (63, 64), further suggest a competition between classical immune suppression and tissue repair by these cells in response to tissue damage in COVID-19 convalescents.

Each Th subset has a paired regulatory subset (28), and this includes Tfh subsets, as B cell help in germinal centres also requires regulation in the steady state (65). In a stereotypical antiviral immune response, Th1 cells migrate to sites of viral infection to establish an adaptive response, and regulatory cells co-migrate to limit chronic inflammation once the pathogen levels decline, however, there is an emerging function of tissue resident Treg cells in tissue repair (62, 66). We did not observe increased Th1 cells, but we did observe a reduction of Th9 cells, which are believed to home to the gut mucosa (67), potentially suggesting a diversion of Th cells to other sites. We also observed that the maturation of Th pools was enhanced in both Th17 and Th22 subsets, where CM marker proportions were increased at all timepoints post-infection. This may suggest that epithelial homing and tissue damage trigger activation and form part of the COVID-19 T cell recall response. It is intriguing that the Treg partners of these lineages, including ThR2, ThR22 and ThR2/22 were all significantly reduced over the same time course post-infection, suggesting that the signal recruiting Th cells to tissue locations are persistent long after COVID infection. A similar imbalance in follicular help vs follicular regulation was also observed, whereby Tfh1 and Tfh2/22 cells were significantly elevated post COVID-19, but total TfhR, TfhR2, TfhR22 and TfhR2/22 cells were reduced. Other studies have demonstrated that CXCR5^+^ Tfh populations are significantly elevated in individuals recovering from COVID-19 and correlate with robust humoral immunity (68), however this previous study did not analyse the regulatory arm in this compartment. Another previous study has reported a decline in Tfh cells at 4 months post-infection (53).

In addition to immunophenotyping by flow cytometry, we performed RNA sequencing of total RNA from 138 blood samples collected from convalescent individuals at 12, 16 and 24 wpi, as well as HCs. To our knowledge, no other study has profiled transcriptome-wide changes in COVID-19 convalescents for such a long period post-infection. We found that the blood transcriptome of convalescents was significantly perturbed compared to HCs, with the largest numbers of DEGs being identified at 12 wpi. Transcriptional dysregulation persisted until at least 24 weeks. There was a very strong enrichment for pathways and BTMs related to transcription, translation, and ribosome biosynthesis among genes up-regulated in recovering individuals, at all 3 timepoints. Many viruses upregulate rRNA synthesis during infection [42, 43], but why rRNA gene expression remains up-regulated months after infection is currently unknown. Other statistically enriched pathways among up-regulated genes included neutrophil degranulation, antimicrobial peptides, immune system and pathways related to other viral infections. These data suggest ongoing inflammatory responses and immune dysregulation in COVID-19 convalescents weeks-to-months after infection. Consistent with these data, neutrophil degranulation has reported to be significantly up-regulated in active infection (69, 70), suggesting that certain signatures of active infection persist well into convalescence.

We also found evidence for dysregulated expression of genes involved in oxidative phosphorylation, a signature which has also been identified in one other recent study of convalescents to occur irrespective of whether elevated inflammatory markers persist or not (20), but whose functional significance is currently unknown. While some changes in gene expression were associated with variation in specific immune cell populations between individuals, differences in gene expression were not solely explained by changes in the frequency of any single immune cell population. A patient-specific analysis of the gene expression activity of pre-annotated BTMs enabled a more thorough assessment of the variation in gene expression responses. There was a broad spectrum in the recovery of gene expression responses in both mild/moderate and severe/critical convalescents. Variation in the rate of recovery from infection at a cellular and transcriptional level may explain the persistence of symptoms, such as fatigue, associated with long COVID in some convalescent individuals, although data related to ongoing symptoms was unfortunately not collected for this cohort. Interestingly, a link between gene expression in peripheral blood and fatigue following infectious mononucleosis has been previously reported (71), with at least some of the same genes differentially expressed in COVID-19 convalescents. These data may point towards common mechanisms regulating ‘long COVID’ and post-viral infection fatigue more generally. Finally, we also uncovered significant inverse correlations between dysregulated BTMs and anti-Spike and anti-RBD antibody responses suggesting that prolonged transcriptional dysregulation may be associated with reduced antibody responses with potential consequences for the durability of protective immunity. Further work is now needed to assess whether dysregulated immunity following COVID-19 has implications for responses to other infections, vaccination or in the management of chronic diseases.

## Limitations of the study

While our study provides a high-resolution, multi-level insight into the immune dysregulation experienced post COVID-19, we recognise that our study also has some important limitations. While comparable to or larger than most other studies to date, the sample size is still relatively limited, particularly in the case of patients with more severe disease. This is particularly important given the apparently highly heterogenous recovery in immune dysregulation over time. Further larger studies will be needed to more fully assess differences due to disease severity, treatment and other confounders. Another limitation is the lack of information on ongoing symptoms experienced by study participants, preventing us from linking specific types of immune dysregulation with particular long COVID symptoms. Other single-cell approaches may also provide further resolution of the immune dysregulation experienced by convalescents. While our flow cytometry analyses enabled the assessment of ∼130 parameters, it did not include markers for dendritic cells (DC), which have been found to be altered in COVID-19 convalescents in previous studies (72). Our BTM analysis, however, supports the dysregulation of DC populations in convalescents. Finally, while we assessed the relationships between immune dysregulation and anti-Spike and anti-RBD antibody responses, we did not assess T cell immunity in our study (73, 74). Further studies should also assess the effects of SARS-CoV-2 variants on long-term immune dysregulation in convalescents and comparative studies assessing differences between post-infectious immune dysregulation following SARS-CoV-2 infection in comparison to other infections would be highly beneficial.

## Methods

### Patient Recruitment

Study participants were recruited via the Central Adelaide Health Network (CALHN). The study was performed in accordance with the ethical principles consistent with the latest version of the Declaration of Helsinki (version Fortaleza 2013), Good Clinical Practice (GCP) and according to the National Health and Medical Research Council (NHMRC) Guidelines for Research published in the National Statement on the Ethical Conduct in Human Research (2007; updated 2018). The protocol was approved CALHN Human Research Ethics Committee, Adelaide, Australia (Approval No. 13050). Inclusion criteria were PCR-confirmed SARS-CoV-2 infection from nasopharyngeal swabs, the ability to attend study follow up visits, and voluntary informed consent. Due to the lack of knowledge of SARS-CoV-2 infection in early-to-mid 2020, the study size was determined in a pragmatic fashion by opportunistically recruiting as many participants as possible. A total of 69 COVID-19 convalescent individuals (35 male, 36 female) representing a range of prior mild, moderate, severe, critical COVID-19 cases were recruited (**Table S1**). COVID-19 disease severity was scored as per NIH descriptors (21) where 5 = “asymptomatic”, 4 = “mild”, 3 = “moderate”, 2 = “severe”, 1 = “critical” (**Table S1)**. Blood samples were collected from convalescents at 12, 16 and 24-weeks (+/- 14 days) post the date of their initial PCR-positive test (which occurred in March & April 2020 for all participants). Participation at each timepoint was determined by availability to attend follow up session. Healthy controls (n=14) in the same ranges of age and sex as the COVID-19 convalescent cohort were also recruited. Healthy controls had no respiratory disease, no positive COVID-19 PCR test in 2020/21, no known significant systemic diseases and negative anti-Spike and anti-RBD serology. Blood (54ml/individual) was collected in serum separator (acid citrate dextrose (ACD)) tubes or ethylenediaminetetraacetic acid (EDTA) tubes and processed for serum, peripheral blood mononuclear cells (PBMCs) and plasma isolation. 2.5 mL of blood for RNA sequencing was collected into PAXgene® tubes (762165 BD, North Ryde, Australia) and stored at -80 °C until processing.

### SARS-CoV-2 PCR testing

Extraction of RNA was achieved from nasopharyngeal swabs using the Automated MagMAX nucleic acid extraction protocol (Thermofisher) and RNA subjected to a one-step qRT-PCR using a Roche light cycler LC408II using cycle conditions described by Corman *et al.* (75).

### SARS-CoV-2 protein purification and ELISA

Prefusion SARS-CoV-2 ectodomain (isolate WHU1, residues1-1208) with HexaPro mutations (76) (kindly provided by Adam Wheatley) and SARS-Cov-2 receptor-binding domain (RBD) with C-terminal His-tag (77) (residues 319-541; kindly provided by Florian Krammer) were overexpressed in Expi293 cells and purified by Ni-NTA affinity and size-exclusion chromatography. Recombinant proteins were analyzed via a standard SDS-PAGE gel to check protein integrity. Gels were stained with Comassie Blue (Invitrogen) for 2 h and de-stained in distilled water overnight. MaxiSorp 96-well plates were coated overnight at 4°C with 5μg/mL of recombinant RBD or S proteins. After blocking with 5% w/v skim milk in 0.05% Tween-20/PBS (PBST) at room temperature, serially diluted (heat inactivated) sera were added and incubated for 2h at room temperature. Plates were washed 4 times with 0.05% PBST and secondary antibodies added. Secondary antibodies were diluted in 5% skim milk in PBST as follows: Goat anti-Human IgG (H+L) Secondary Antibody, HRP (1:30,000; Invitrogen); Mouse Anti-Human IgG1 Fc-HRP (1:5000, Southern Biotech), Mouse Anti-Human IgG3 Hinge-HRP (1:5,000; Southern Biotech); goat anti-human IgM HRP (1:5,000; Sigma): anti-human IgA HRP antibody (1:5,000; Sigma) and incubated for 1 hour at room temperature. Plates were developed with 1-Step™ Ultra TMB Substrate (Thermo Fisher) and stopped with 2M sulphuric acid. OD readings were read at 450nm on a Synergy HTX Multi- Mode Microplate Reader. AUC calculation was performed using Prism GraphPad, where the X-axis is half log10 of sera dilution against OD450 on Y-axis.

### PBMC isolation

Post plasma centrifugation, the white blood cell pack was harvested, pooled into 1x 50ml falcon tube, diluted in 2% FCS/PBS up to 35ml and overlayed onto 15ml Ficoll, centrifuged for 20 minutes, 1000 x *g*, RT, no brake. The PBMC were isolated, washed in 2% FCS/PBS, centrifuged at 480 x *g* for 10 minutes at RT, PBMC resuspended in 50ml 2% FCS/PBS, manually counted using trypan blue exclusion assay, 2×10^6^ cells were plated across 4 wells (5×10^5^ per well) of a 96 well plate. The 50ml tube was then spun at 300 x *g* for 10 minutes, the pellet was resuspended in ½ volume of FCS with ½ volume of 20% DMSO/80% FCS added dropwise to final cell concentration of 1×10^7^ per ml. The samples were stored 800µL- 1.8ml per vial placed in a CoolCell at -80°C. The frozen PBMC tubes were transferred to liquid nitrogen for long-term storage within 1-7 days.

### Flow cytometry staining

The 96 well plate was centrifuged at 300 x *g* for 4 mins, the plate was inverted on paper towel and the PBMC pellets were stained with 30µL of 1 of 3 master-mixes of antibodies (lineage, 15 color; memory, 8 color; T helper/Treg, 14 color) for 20 minutes RT, in dark, which included a co-stain of BD LIVE/DEAD fixable dye (stained at 1:1000). The stained PBMC were washed with 200µL of FACS wash, centrifuged 300 x *g* for 4 minutes and fixed with 200µL FACS Fix for 20 minutes, RT, in dark. Fixed cells where then centrifuged 300 x *g* for 4 mins, washed in 200µL FACS wash then spun 300 x *g* for 4 mins and resuspended in 50µL FACS wash. The cells were resuspended and added to tubes before being analyzed using a BD FACS Symphony within 3 days of staining/fixing.

### Flow cytometry data acquisition & analysis

To control for batch effects, the BD FACS symphony lasers are calibrated with dye conjugated standards (Cytometer Set &Track beads) run every day. All samples were acquired with all 28 PMTs recording events. All voltages of PMTs were adjusted to negative unstained control baseline typically log scale 10^2^. Antibodies were titrated for optimal signal over background so that single positive stains sat within log scale 10^3^-10^5^ of designated PMT. Compensation was set with beads matched to each panel antibody combination using spectral compensation using FlowJo Software V10. Exported FCS files had compensation values adjusted manually post-acquisition on a file-by-file basis in FCS express v6. Once compensated, low data quality events were excluded based upon time acquired (at the sample acquisition start and before sample exhaustion), with further time exclusion gates based on blockages or unexplained loss of events for a period of time during acquisition. Events that were highly positive for LIVE/DEAD staining were removed from subsequent analysis, to prevent exclusion of live monocytes, which take up more live/dead dye per cell than T cells, giving a high background. Events were gated for FCS-H/A as well as SSC-H/A linearity, and restricted FSC-W and SSC-W values for doublet discrimination. Live single cells were then broadcast on SSC-A / FSC-A plot to determine size and complexity. Lymphocyte, monocyte, and granulocyte gates were based on physical parameters (unless a lineage specific antibody was added). Populations of cells were expressed as a proportion of the highest order lineage gate (namely: Lymphocytes, Monocytes and Granulocytes). For parameters measured in healthy controls and COVID-19 samples at 12 weeks, 16 weeks and 24 weeks, a Wilcoxon Rank Sum test was used to assess statistical significance with the Benjamini and Hochberg method employed to correct for multiple comparisons. Statistical significance was determined as FDR < 0.05.

### RNA extraction and library preparation

RNA extraction and genomic DNA elimination was carried out using the PAXgene® Blood RNA kit (762164, Qiagen, Feldbachstrasse, Germany) as per the manufacturer’s instructions. Final elution into 80 μL RNase-free water. A further RNA precipitation reaction was carried out. Briefly, RNA was resuspended 2.5 x 100% ethanol and 10% sodium acetate and spun at 12,000 x *g* for 30 min at 4C. Samples were washed in 75% ethanol. Pellets were air dried and resuspended in 40 μL RNase free water and total RNA yield was determined by analysis of samples using a TapeStation (Agilent) and Qubit (ThermoFisher Scientific, Australia). Total RNA was converted to strand-specific Illumina compatible sequencing libraries using the Nugen Universal Plus Total RNA-Seq library kit from Tecan (Mannedorf, Switzerland) as per the manufacturer’s instructions (MO1523 v2) using 12 cycles of PCR amplification for the final libraries. An Anydeplete probe mix targeting both human ribosomal and adult globin transcripts (HBA1, HBA2, HBB, HBD) was used to deplete these transcripts. Sequencing of the library pool (2×150 bp paired-end reads) was performed using 2 lanes of an S4 flowcell on an Illumina Novaseq 6000.

### RNA-Seq analysis

Sequence read quality was assessed using FastQC version 0.11.4 (78) and summarised with MultiQC version 1.8 (79) prior to quality control with Trimmomatic version 0.38 (80) with a window size of 4 nucleotides and an average quality score of 25. Following this, reads which were <50LJnucleotides after trimming were discarded. Reads that passed all quality control steps were then aligned to the mouse genome (GRCh38 assembly) using HISAT2 version 2.1.0 (81). The gene count matrix was generated with FeatureCounts version 1.5.0-p2 (82) using the union model with Ensembl version 101 annotation. The count matrix was then imported into R version 4.0.3 for further analysis and visualisation in ggplot2 v2.3.3. Counts were normalized using the trimmed mean of M values (TMM) method in EdgeR version 3.32 and represented as counts per million (cpm) (83). Prior to multidimensional scaling analysis and generation of heatmaps, svaseq v3.38 was applied to remove batch effects and other unwanted sources of variation in the data (84). Differential gene expression analysis was performed using the glmLRT function in EdgeR adjusting for sex and batch (run) in the model. Genes with <3 cpm in at least 15 samples were excluded from the differential expression analysis. Pathway and Gene Ontology (GO) overrepresentation analysis was carried out in R using a hypergeometric test. To assess if differential gene expression was primarily driven by differences in the proportion of any major immune cell population (i.e. LD granulocytes, LD neutrophils, CXCR3^+^ neutrophils, monocytes, lymphocytes, CD56^++^ NK cells, CD19^+^ B cells, CD3^+^ T cells, NKT cells, CD4^+^ T cells or CD8^+^ T cells), we additionally fit the frequency of each population in each individual into the EdgeR model and reperformed the differential gene expression and pathway overrepresentation analysis. Blood Transcriptional Module (BTM) analysis was carried using a pre-defined set of modules defined by Li *et al.* as an alternative to pathway-based analyses (43). Gene Set Variation Analysis (GSVA) (44) was used to calculate a per sample activity score for each of the modules (excluding unannotated modules labelled as ‘TBA’). *limma* v3.46.0 was used to identify modules that were differentially active in at least one timepoint. Pearson correlation analysis was performed using the Hmisc v4.4-2 package in R to determine correlations between anti-Spike and anti-RBD antibody titres, flow cytometry data and BTM activity scores. Correlation networks were exported to Cytoscape v3.8.1 for visualisation.

## Supporting information

Document S1 - Gating strategy for flow cytometry

Supplementary cytoscape correlation network files

STROBE Checklist

## Data Availability

RNA-Seq data have been deposited in the Gene Expression Omnibus (GEO) under accession GSE169687. Count tables, metadata, and R code for all analyses (serology, flow cytometry and RNA-Seq) have been uploaded to the Lynn Laboratory BitBucket (https://bitbucket.org/lynnlab/covid-sa).

## Author contributions

The study was conceived and designed by B.G.B, S.C.B and D.J.L.; C.M.H. and S.C.B. generated the flow cytometry data. F.J.R. performed the RNAseq analysis and the majority of statistical analyses and visualisations in the paper under the supervision of D.J.L. M.A.L. extracted the RNA for RNAseq. A.E.L.Y., M.G.M., Z.A.M., P.G-V., Z.A-D., and B.G.B. generated the serology data. J.G. and C.M.H. sample collection and processing. C.F. S.O’C., B.A.J.R., D.S., C.K.-L., J. G., M.R.B. patient enrolment and management, sample collection. The manuscript was written by F.J.R., B.G.B., S.C.B. and D.J.L with contributions and approval from all of the authors.

## Acknowledgements

We would like to thank the South Australian Genomics Centre for their invaluable assistance with generating the total RNA sequencing data and Natalie Stevens for assistance with data analysis and interpretation.

## Figure and Table Legends

**Document S1:** Flow cytometry gating strategy.

**Figure S1:** Stability of anti-Spike and anti-RBD antibody titres over time. **(A-E)** Anti-Spike and (F-J) anti-RBD IgG, IgG1, IgG3, IgM and IgA titres plotted as a function of time. End point titers are reported as log_10_ area under the curve (AUC). The blue line represents the line of best fit from a linear regression analysis. The shaded areas represent the 95% confidence interval. The P value shown is from the linear regression. Red dashed lines represent the mean AUC + 2 SD in healthy controls for each isotype.

**Figure S2:** Expression of genes in two pathways identified as down-regulated in COVID-19 convalescents and healthy controls. **(A)** Reactome pathway R-HSA-76002 ‘Platelet activation, signaling and aggregation’ and **(B)** KEGG pathway hsa00190 ‘Oxidative phosphorylation’. Oxidative phosphorylation genes are sub-divided into nuclear and mitochondrially encoded, with the same x axis order of samples in each panel. Only differentially expressed genes (FDR < 0.05 and fold change >1.25-fold) within each pathway are shown.

**Figure S3:** Adjusting for differences in immune cell populations. We repeated the differential expression analysis multiple times, each time adjusting for differences in the frequency of major immune cell populations among individuals. Each panel shows the enrichment of selected pathways (same as those shown in Fig. 4F) among genes identified as being significantly up-regulated in each analysis (FDR < 0.05 and fold change > 1.25-fold). None = no immune cell population adjustment.

**Table S1.** Subject meta-data for COVID-19 convalescents and healthy controls and STROBE checklist for reporting a cohort study.

**Table S2.** Flow cytometry data showing the fold-change and FDR values for each immune cell population in COVID-19 convalescents compared to healthy controls.

**Table S3.** Pearson correlations between the frequency of immune cell populations at 12, 16, and 24 weeks post-infection (wpi) and anti-Spike and anti-RBD antibody responses.

**Table S4.** RNAseq sample statistics, differentially expressed genes (DEGs), pathway analyses, and adjusting for immune cell populations.

**Supplementary File 1.** Cytoscape network file related to Figure 5B.

