## Supplementary material for "Long-term perturbation of the peripheral immune system months after SARS-CoV-2 infection": Document S1 - Gating strategy for flow cytometry

- **Flow cytometry gating strategy for; Lymphocytes, Monocytes and Low Density (LD) Neutrophils:** (A) Events over time are gated to remove fluctuations at the start and end of sample acquisition. (B) Live cells are negative for LIVE/DEAD stain. The live events are then subjected to linear gates of both (C) FSC and (D) SSC height and area. The remaining events undergo doublet removal using the width parameters of (E) FSC and (F) SSC. (G-I) Cells are then physically separated using FSC-A and SSC-A physical parameters (H) CD14 MFI and (I) CD16 MFI help guide monocyte and LD neutrophil population gating, respectively. (J) CD16+ LD Neutrophils and (K) CD14+ Monocyte populations were gated using CD14 vs CD16 plots.

- **Gating strategy for Neutrophil, Monocyte, T cell, NKT, NK and B cell subtyping**; (A) CD14 and CD16 expression on LD granulocytes, LD Neutrophils (CD14+ CD16-) (B) CXCR3 and HLA-DR measurement on LD Neutrophils (C) CD19+ B cells gated on CD27 and CD38, Naïve B cells (CD27-CD38+) Memory B cells (CD27+) and CD38+ Memory B cells (CD27+CD38+) where measured. (D) Memory B cells gated on CD24 and CD38 showing outlined Transitioning B cell gate. (E) Naïve B cells gated on CD24 and CD38 with events gated for CD24+CD38++ Transitioning B cells. (F) Monocyte subtyping using CD14 and CD16: CD14+CD16- Classical, CD14+CD16+ Intermediate and CD14-CD16- non-classical. (G) CD14+ Monocytes CXCR3 and HLA-DR status. (H) CD3 and CD19 used to define; B cells (CD19+), T cells (CD3+), and nBnT (CD19-CD3-) lymphocytes. (I) CD4 and CD8 staining of T cells defining CD4 T cells, CD8 T cells, CD4+CD8+ Double Positive and CD4-CD8- Double Negative T cells (J) CD56 and CD16 expression on T cells, gating for CD56+NKT and CD16+NKT. (K) Non-B and Non-T cells (nBnT) population showing expression of CD56 and CD16 to identify CD56Bright (CD56++), NK cells (CD56+CD16+) and CD56-CD16+ NK cells. (L-R) CXCR3 and HLA-DR expression on; (L) CD4 T cells (M) CD8 T cells (N) CD56+ T cells (O) CD16+ T cells (P) CD56++ NK cells (Q) CD56+CD16+ NK cells (R) CD56-CD16+ NK cells. (S-W) CD27 and CD38 expression on; (S)CD4-CD8- DN T cells (T) CD4 T cells (U) CD8 T cells (V) CD56+ T cells (W) CD16+ T cells. CD4 and CD8 expression on (X) CD56+ T cells (Y) CD16+ T cells.

- **Flow cytometry gating strategy for CD4 and CD8 Naïve and memory T cell subsets**; (A) CD3 used to identify T cells (B) CD4 and CD8 used for T cell subsets. CD45RA and CD45RO isoforms used to initially gate CD45RA+CD45RO-,CD45RA-CD45RO+ as well as events that do not lie within those gates CD45RAloCD45ROlo Transitioning memory T cell subsets in (C) CD4 T cells and (D) CD8 T cells. (E-G) CD4 T cell subsets expressing (E) CD45RA+CD45RO- (RO-), (F) Transitioning memory and (G) CD45RA- CD45RO+ (45RO+). (H-J) are the CD8 T cell subsets expressing (H) CD45RA+ CD45RO- (RO-), (I) Transitioning memory and (J) CD45RA- CD45RO+ (RO+). (K) Gating hierarchy. Using this gating strategy there are 24 populations investigated. Central Memory (CM) are CCR7+, with loss of CCR7 indicating an Effector Memory (EM) population. “Loss” of CD62L from CM was denoted with a (-) sign. “Gain” of CD62L on EM denoted with (+) sign.

- **Flow cytometry gating strategy for CD4 T follicular helpers (Tfh), T helpers (Th), T follicular Tregs (TfR), and T helper Tregs (ThR);** (A) CD4 and CD8 utilised to identify T cell pools, (B) CD127 and CD25 used for gating Conventional T cells (Tconv, CD127+ CD25mid) and Tregs (CD127lo + CD25hi). CD45RO and CD62L were used to identify naïve (CD45RO- CD62L+) and memory (CD45RO+) status for (C) Tconv and (D) Tregs. CXCR5 and CD45RO were used to determine (E) T follicular helpers (Tfh, CXCR5+CD45RO+) and T helpers (Th, CXCR5-CD45RO+) as well as (F) T follicular Tregs (TfR, CXCR5+CD45RO+) and T helper Tregs (ThR, CXCR5-CD45RO+). PD-1 and ICOS were used as activation markers of (G) Tfh and (H) TfR subsets.

- **Chemokine Receptor gating strategy for CD4 T follicular helpers (Tfh), T helpers (Th), T follicular Tregs (TfR), and T helper Tregs (ThR);** (A)CXCR3 and CCR6 used to determine Th1 (CXCR3+CCR6-), Th1/17 (CXCR3+CCR6+). CXCR3-CCR6+ (blue shaded) and CXCR3-CCR6- (orange shaded) populations used in subsequent gating. (B) CCR4 and CCR10 were used to split the CXCR3-CCR6+ population into Th9 (CCR4-CCR10-), Th17 (CCR4+CCR10-) and Th22 (CCR4+CCR10+). (C)CCR4 and CCR10 were used to split the CXCR3-CCR6- population into ThNEG (CCR4-CCR10-), Th2 (CCR4+CCR10-) and Th2/22 (CCR4+CCR10+). This gating strategy was used on (D-F) CXCR5+CD45RO+ Tfh, (G-I) CXCR5-CD45RO+ Th, (J-L) CXCR5+CD45RO+ TfhR and (M-O) CXCR5-CD45RO+ ThR.

### Gating strategy for Lymphocytes, Monocytes and Low Density (LD) Neutrophils

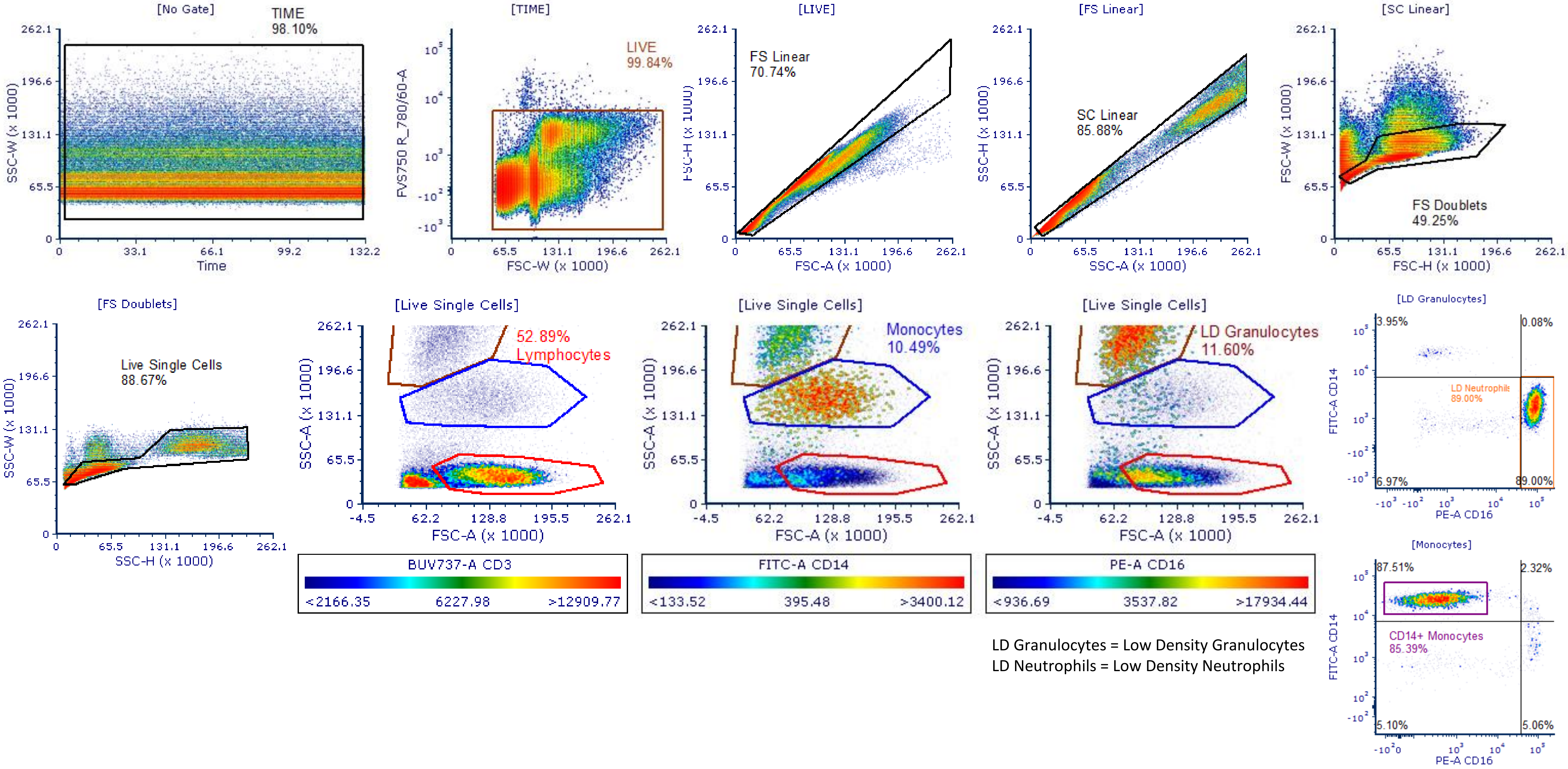


LD Granulocytes = Low Density Granulocytes
LD Neutrophils = Low Density Neutrophils

### Gating strategy for Neutrophil, Monocyte, T cell, NKT, NK and B cell subtyping

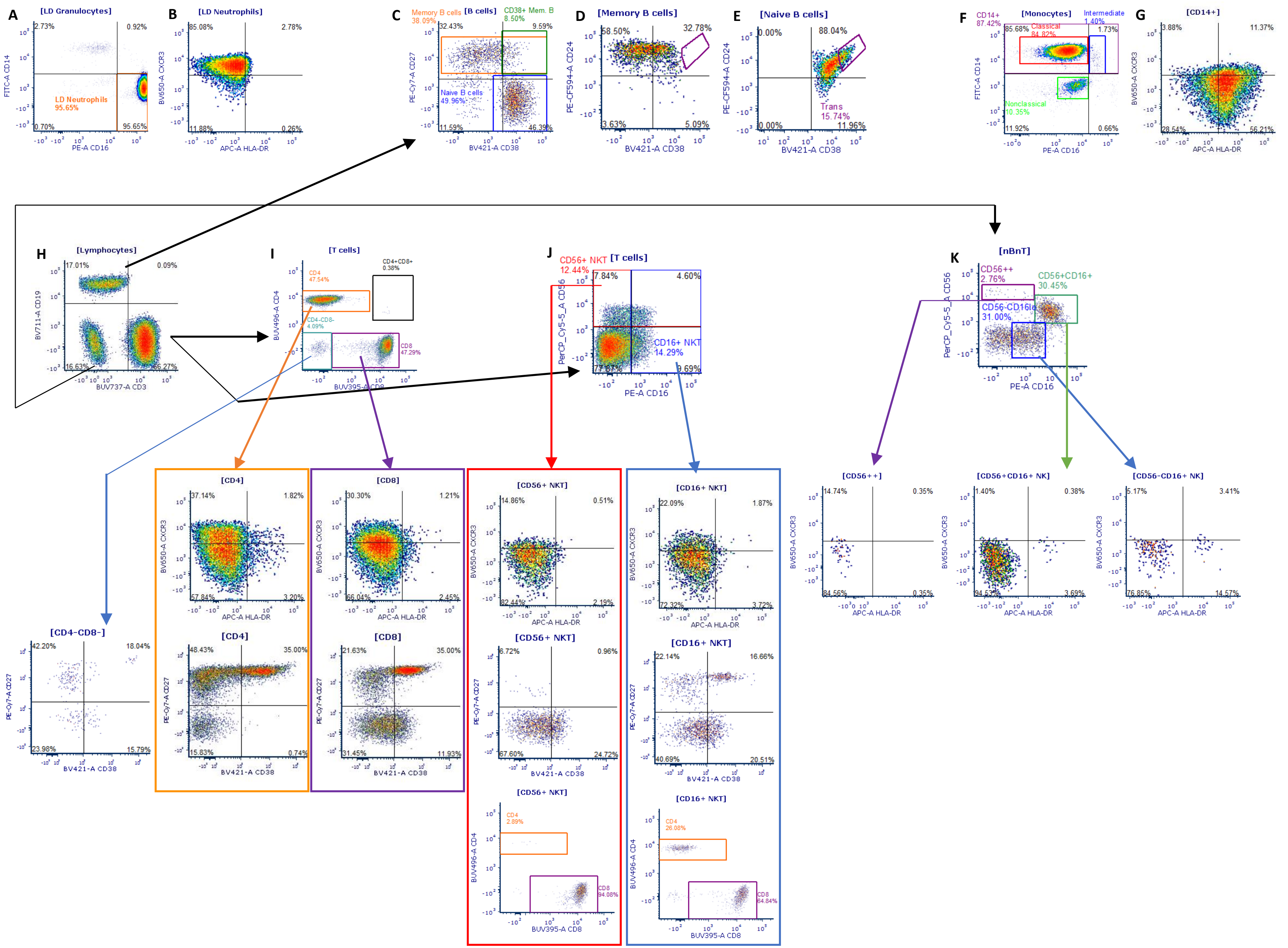

### Gating strategy for CD4 and CD8 Naïve and memory T cell subsets

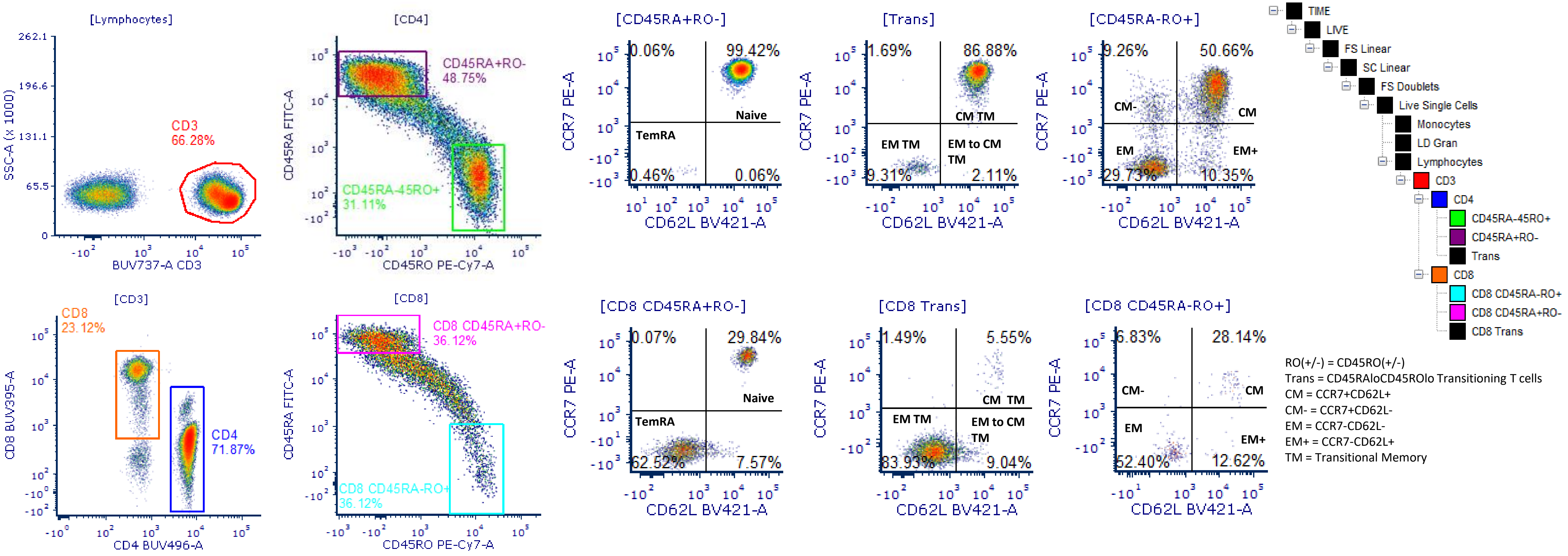


RO(+/-) = CD45RO(+/-)
Trans = CD45RAloCD45ROlo Transitioning T cells
CM = CCR7+CD62L+
CM- = CCR7+CD62L-
EM = CCR7-CD62L-
EM+ = CCR7-CD62L+
TM = Transitional Memory

### Gating strategy for CD4 T follicular helpers (Tfh), T helpers (Th), T follicular Tregs (TfR), and T helper Tregs (ThR)

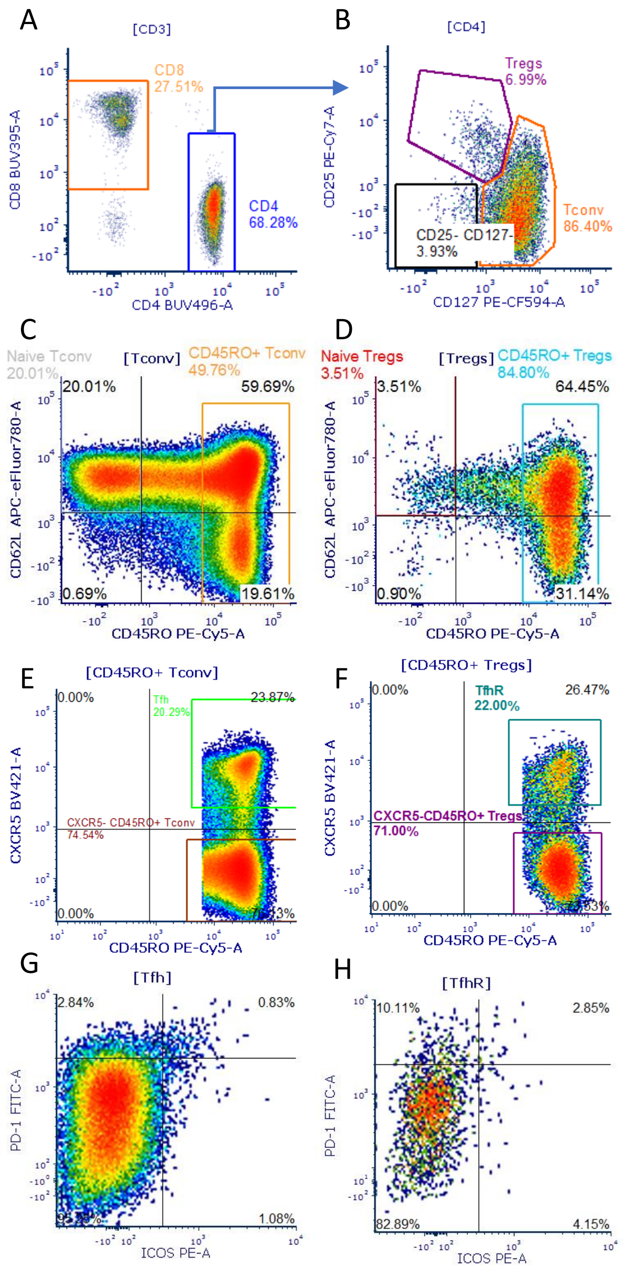

Chemokine Receptor gating strategy for CD4 T follicular helpers (Tfh), T helpers (Th), T follicular Tregs (TfR), and T helper Tregs (ThR);

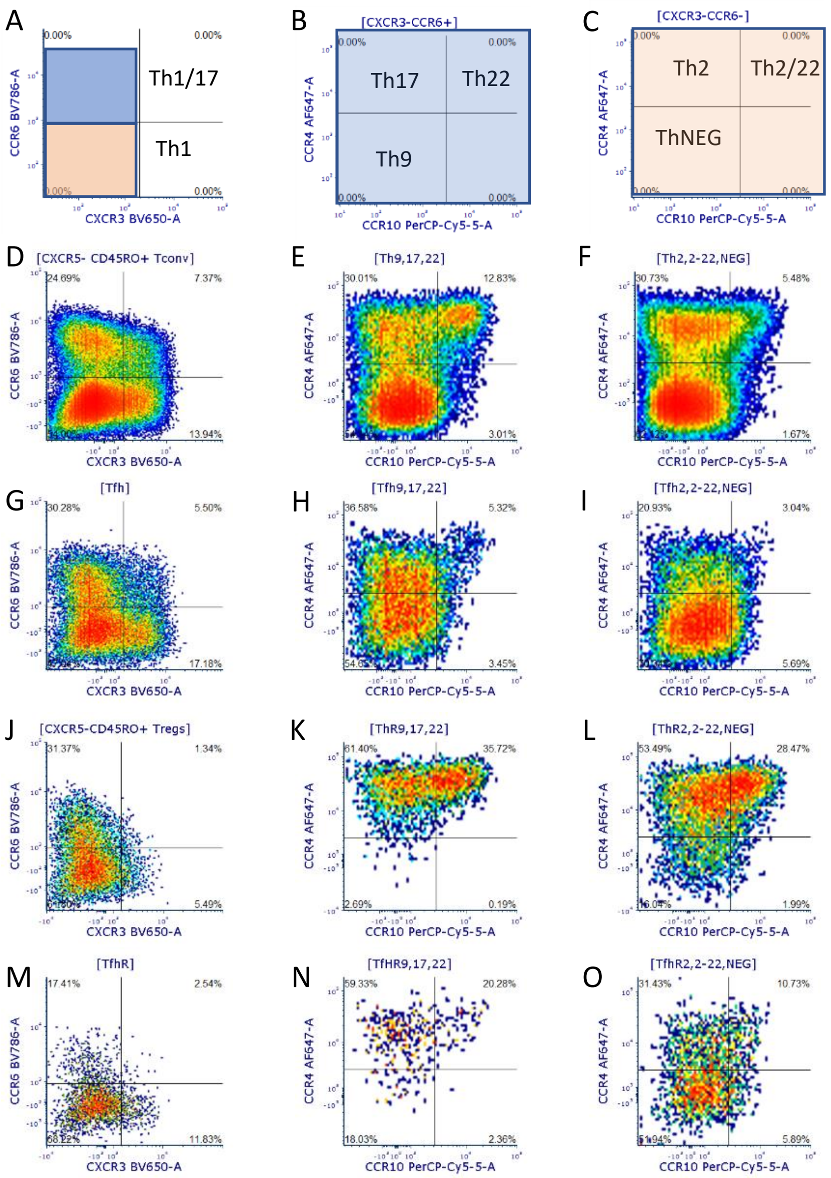
