## Supplementary figures and images for "Long-term perturbation of the peripheral immune system months after SARS-CoV-2 infection"

### Figure S1

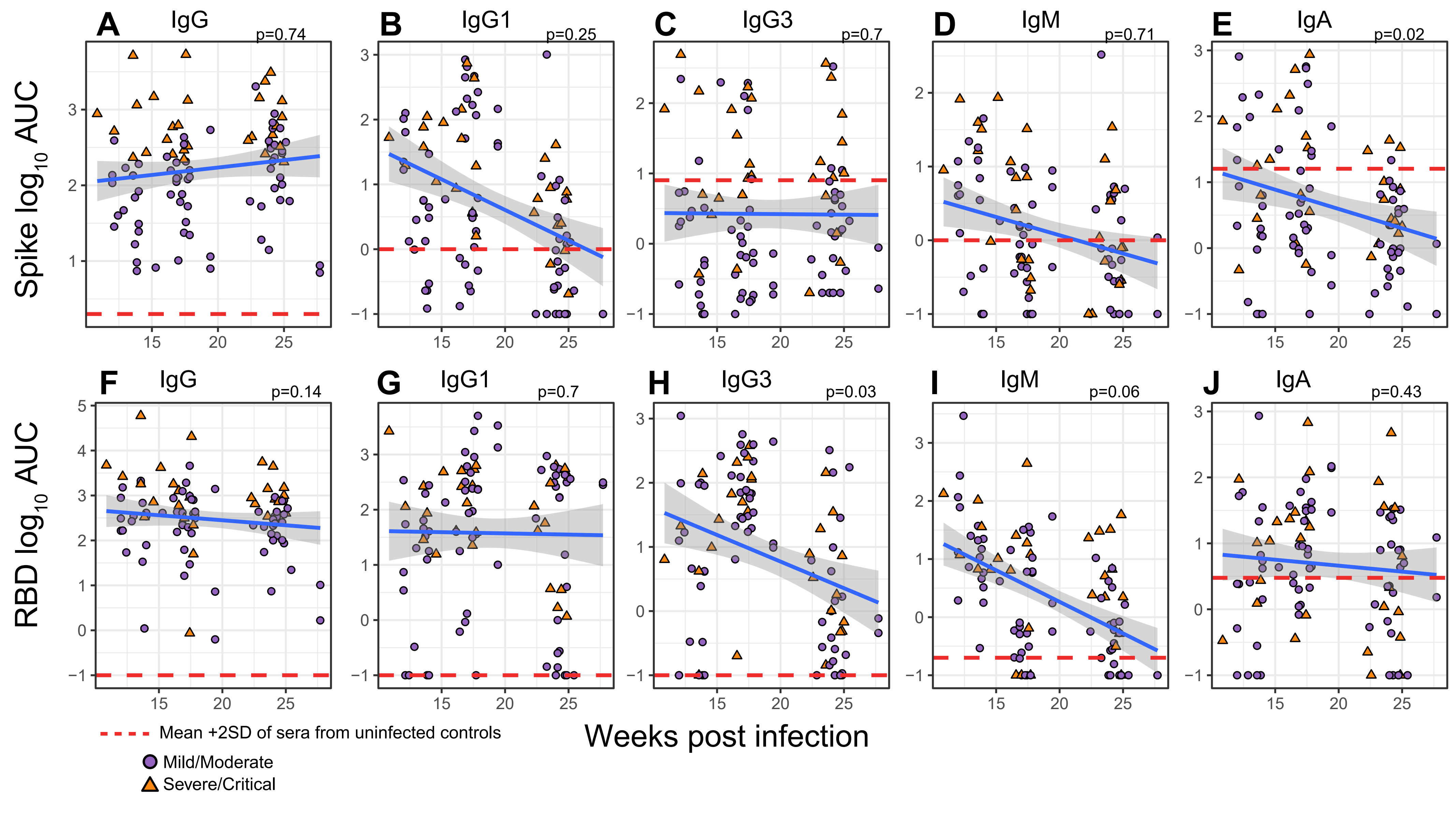

### Figure S2

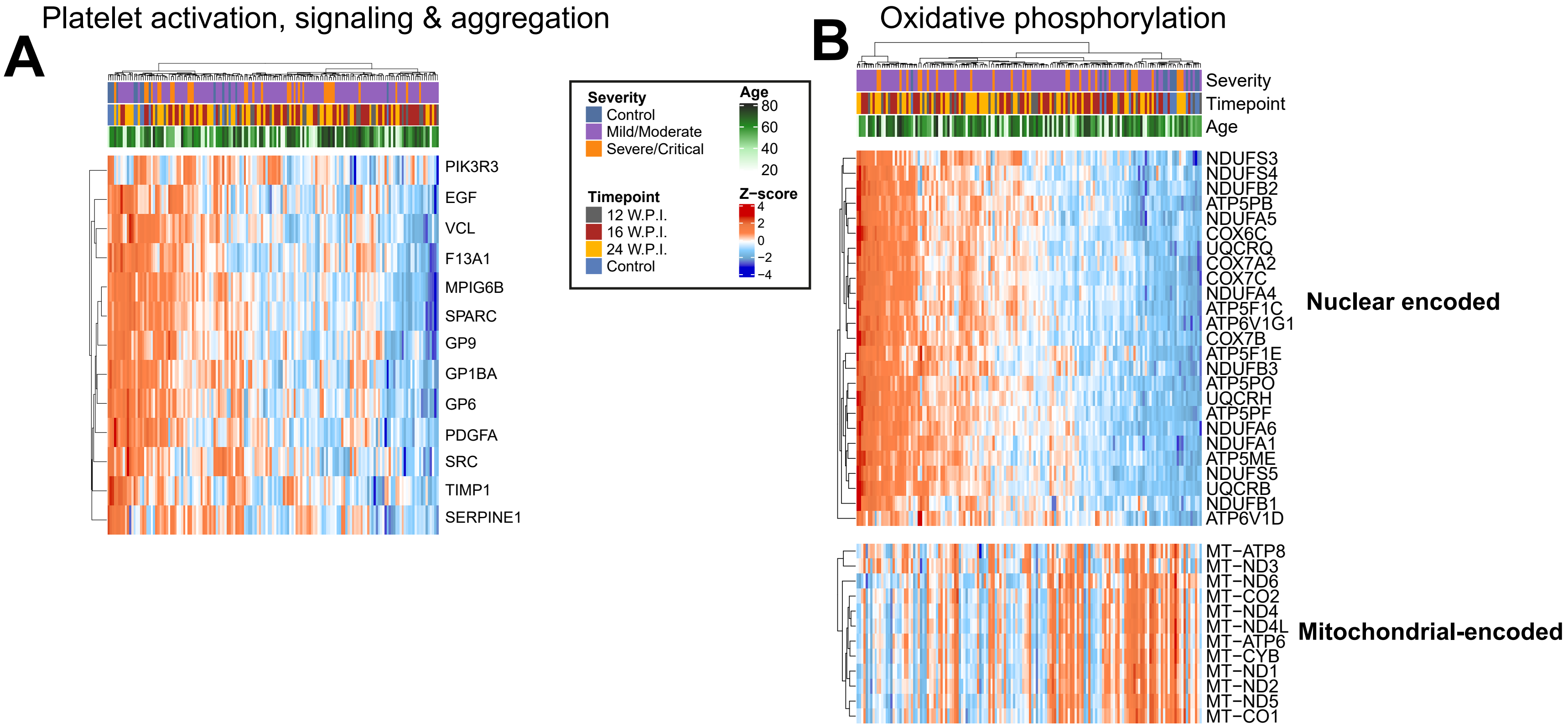

### Figure S3

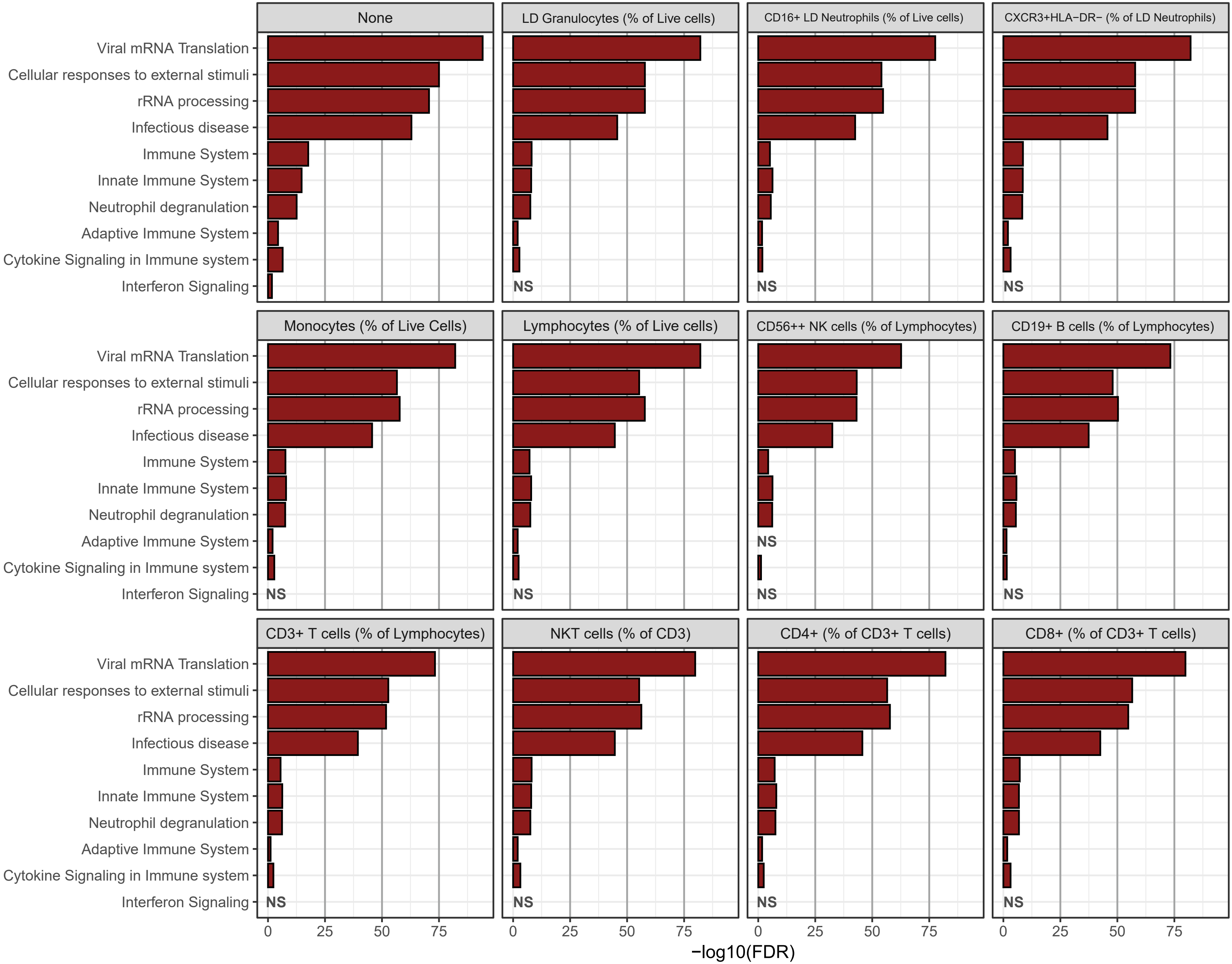
